# The genetic diversity of Nipah virus across spatial scales

**DOI:** 10.1101/2023.07.14.23292668

**Authors:** Oscar Cortés Azuero, Noémie Lefrancq, Birgit Nikolay, Clifton McKee, Julien Cappelle, Vibol Hul, Tey Putita Ou, Thavry Hoem, Philippe Lemey, Mohammed Ziaur Rahman, Ausraful Islam, Emily S. Gurley, Veasna Duong, Henrik Salje

## Abstract

Nipah virus (NiV), a highly lethal virus in humans, circulates silently in *Pteropus* bats throughout South and Southeast Asia. Difficulty in obtaining genomes from bats means we have a poor understanding of NiV diversity, including how many lineages circulate within a roost and the spread of NiV over increasing spatial scales. Here we develop phylogenetic approaches applied to the most comprehensive collection of genomes to date (N=257, 175 from bats, 73 from humans) from six countries over 22 years (1999–2020). In Bangladesh, where most human infections occur, we find evidence of increased spillover risk from one of the two co-circulating sublineages. We divide the four major NiV sublineages into 15 genetic clusters (emerged 20-44 years ago). Within any bat roost, there are an average of 2.4 co-circulating genetic clusters, rising to 5.5 clusters at areas of 1,500-2,000 km^2^. Using Approximate Bayesian Computation fit to a spatial signature of viral diversity, we estimate that each genetic cluster occupies an average area of 1.3 million km^2^ (95%CI: 0.6-2.3 million), with 14 clusters in an area of 100,000 km^2^ (95%CI: 6-24). In the few sites in Bangladesh and Cambodia where genomic surveillance has been concentrated, we estimate that most of the genetic clusters have been identified, but only ∼15% of overall NiV diversity has been uncovered. Our findings are consistent with entrenched co-circulation of distinct lineages, even within individual roosts, coupled with slow migration over larger spatial scales.

## Introduction

Nipah virus (NiV) is a bat-borne zoonotic virus, classified by the World Health Organization as a priority pathogen (*1*). The majority of infected humans die following infection and while most infections occur following zoonotic spillover events, human-to-human transmission is responsible for around a third of known cases (*2*). There is currently no available treatment or vaccine. NiV was first identified in 1999 in Malaysia and has since been detected almost annually throughout South Asia (*3–5*). *Pteropus* fruit bats are the reservoir hosts for NiV (*6*). Detected spillovers in humans primarily occur when individuals drink raw date palm sap from trees upon which infected bats have fed (*4*). However, infection through intermediary species, such as pigs in the 1999 outbreak in Malaysia and horses in 2014 in the Philippines, has also been registered (*4, 7*).

Despite the substantial risk to human health, very little is known about the underlying genetic diversity of NiV. We do know that *Pteropus* bats are found throughout South and Southeast Asia and that NiV infection in *Pteropus* bats is common, with serostudies from across the region identifying 3-83% of adult bats having antibodies to NiV (*3, 8, 9*). However, the patterns of infection within bat populations remain unclear, including the number of discrete lineages circulating within any roost, the extent to which different lineages spread through the region or the ability for NiV to freely transmit between different species within the *Pteropus* genus. We also do not know if specific lineages are linked to increased spillover risk, including the specific routes of transmission. These critical knowledge gaps are problematic for assessments of spillover risk, and vaccine development.

Characterizing the genetic diversity of NiV is difficult as many isolates of NiV are from human cases, which remain rare events and are only detected in a few places. Even in Bangladesh, the country with most spillover events, there are only estimated to be ∼15 spillovers into humans per year (*10*). The representativeness of viruses obtained from human infections is also unclear. This has motivated efforts to instead obtain NiV from bat populations, but this is very difficult as bats appear to be largely asymptomatic and virus shedding is detected infrequently (*9*). These efforts have also been concentrated to a few locations within Thailand, Bangladesh and Cambodia (*11–13*). While it is difficult to make inferences about NiV diversity using sequences obtained from any one location, by pooling information across these different locations combined with appropriate analytical methods, we can obtain a more complete picture of diversity. Here we provide a comprehensive assessment of the diversity of NiV using all publicly available sequences coming from six countries, along with several previously unpublished sequences. We develop methods that are robust to strong biases in where and when sequences are obtained to track the diversity of NiV across spatial scales (within roost, within district, country and international) and quantify the extent to which diversity has been fully identified in locations that have implemented extensive virus surveillance efforts.

## Results

We analyzed 257 sequences from six countries covering a 22-year time period (1999-2020, Figure 1A, Figure S1). 73 (28%) of the sequences came from human infections, 175 (68%) from bats and five (4%) from other sources (Figure 1B). Using Bayesian Evaluation of Temporal Signal (BETS) analysis (*14*), we found there was strong support for a model in which the sampling dates of the sequences were included over a model in which they were considered contemporaneous (Bayes Factor of 415). We therefore built a time-resolved phylogeny and found that sequences could be broadly characterized into two major genotypes: genotype I and genotype II, previously reported (*15*), and four minor genotypes I.A, I.B, II.A, II.B (Figure 1C). We estimated a mean nucleotide substitution rate of 4.5 x 10^-4^ subs/site/year (95%CI: 2.9 x 10^-4^, 6.0 x 10^-4^), consistent with previous estimates (*12*). We found that genotypes I and II diverged in 1937 (95%CI: 1838-1983), and the minor genotypes diverged in the 1970s. There was broad spatial structure in these genotypes, with genotype I being dominant in the west of the region and genotype II in the east of the region (Figure 1D). We found that countries on the Eastern (Indonesia/Malaysia) and Western edges (India) only had a single circulating sublineage. By contrast, Cambodia, with a central position in the region, had sequences from all four sublineages.

**Figure 1:**
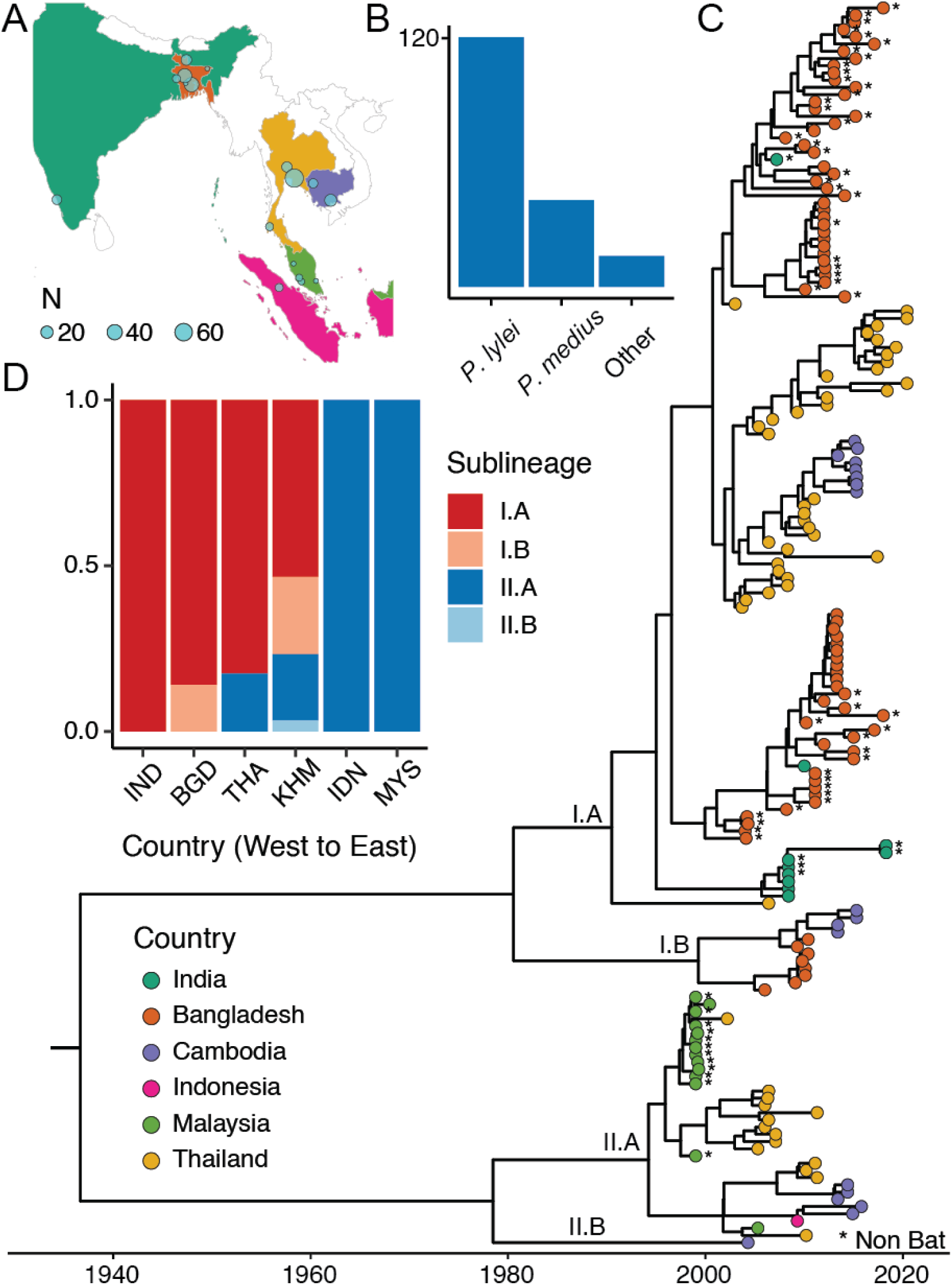
(A) Country of origin sequences. **(B)** Distribution of host species for sequences coming from bats (N=175). **(C)** Reconstructed time resolved Maximum Clade Credibility phylogeny with tips coloured with country of origin and non-bat sequences are marked with an asterisk. **(D)** Sublineage distribution per country.

Three quarters of human sequences obtained from human cases were from Bangladesh (N=55/73), which had two circulating sublineages (IA and IB). Lineage IA was found throughout the period 2004-2018, whereas IB was found in the period 2006-2009 only (Figure S2). We found that all human sequences in Bangladesh (N=55/55) came from genotype IA, with none from genotype IB (N=0/55). By contrast, 64.9% (N=24/37) of the bat sequences come from IA and 35.1% (N=13/37) come from IB. These observations suggest that some genotypes may have an increased probability of spilling over into human populations (p-value of 0.007 using Fisher exact test).

To assess the extent of spatial structure among the sequences, we used the time-resolved phylogeny to compare the total evolutionary time that separated each pair of viruses with their spatial separation. To limit the effects of sampling bias from the sequences obtained from bat roosts we randomly sampled a single sequence from each roost and each year, and we sampled a single sequence from each human case cluster. We found that each 10-year increase in evolutionary time was associated with an average 186 km (95% CI 161 - 212) increase in the distance that separates them, equivalent to 19 km a year (Figure 2).

**Figure 2:**
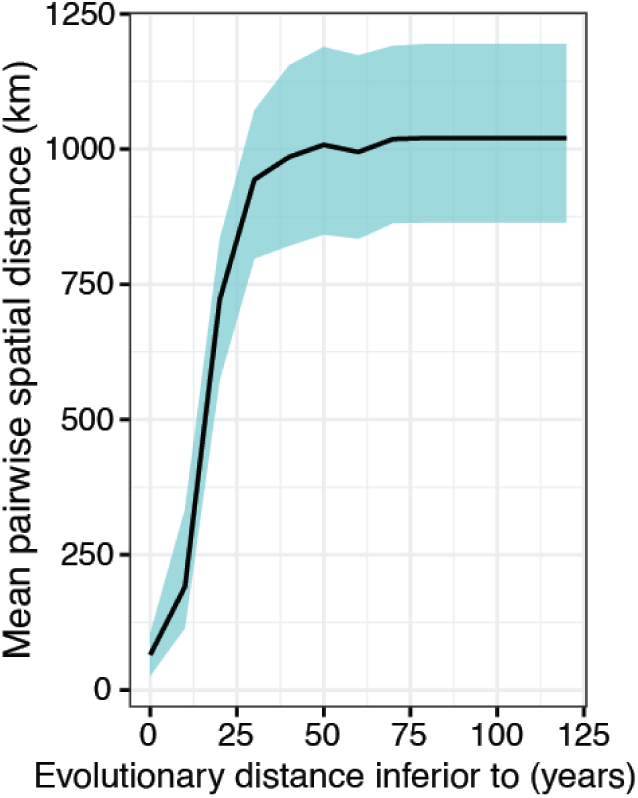
Mean pairwise spatial distance (in km) in function of pairwise evolutionary distance (in years).

In order to provide a finer scale characterization of genetic diversity, we adapted a genetic clustering method, PhyCLIP, to sort the sequences into different genetic clusters based on the distribution of evolutionary distances between pairs in the phylogeny (*16*). This approach resulted in 15 unique clusters (Figure 3A-D). To assess the robustness of our cluster assignments, we reimplemented the clustering algorithm on 100 randomly selected posterior trees. We found highly consistent grouping of sequences (median Adjusted Rand Index of 0.94, where a value of 0 would indicate random group assignment and 1.0 perfect consistency in group assignment) (*17*).The dates of divergence of the genetic clusters ranged from 1978 to 2002, with the mean evolutionary time from a cluster to the next-closest cluster of 15 years. We found that genetic clusters tended to aggregate within the same country (Figure 3A). The clusters were also separated by bat species with eleven of the genetic clusters exclusively found within one species (Figure 3B). Bat species themselves were also spatially structured at a country level, as all of the sequences from four (67%) countries came from one single bat species (Figure 3C). We estimated that 96.8% (95%CI: 96.4-97.2) of bat roosts separated by <100 km are of the same species, dropping to 53.4% (95%CI 47.9-58.8) for roosts separated by 500-1,000 km (Figure S3). On average, each 100 km increase in distance between bat roosts is associated with 0.62 (95%CI 0.61 - 0.63) times the odds of the roosts coming from the same species, providing a crude quantification in the spatial overlap of bat species in the region.

**Figure 3:**
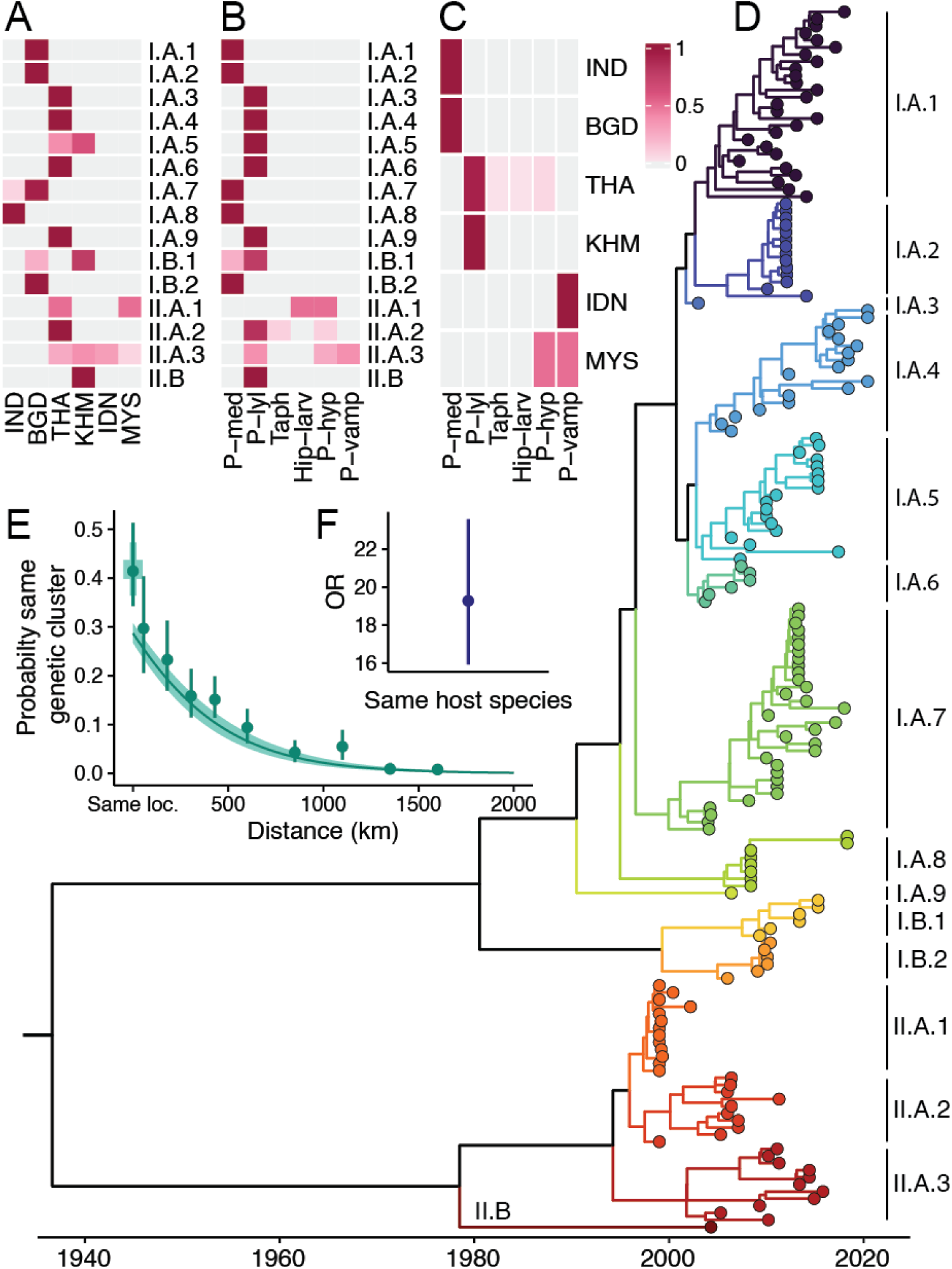
(A) Spatial distribution of identified lineages for bats. Each line represents the proportion of sequences coming from a specific country for the corresponding genetic cluster. **(B)** Bat host species distribution of lineages. **(C)** Distribution of bat host species per country. **(D)** Time-resolved phylogeny divided into fifteen distinct clades using an adapted form of PhyCLIP. **(E)** Proportion of sequence pairs belonging to the same cluster as a function of their spatial distance. The green shaded region represents 95% confidence intervals. **(F)** Odds ratio of belonging to the same genetic cluster if sequence pairs were sampled from the same bat host species or not. The error bar represents 95% confidence intervals.

We found that there were an average of 2.41 (95%CI: 1.92-2.94) different genetic clusters found within the same bat roost. The probability that a pair of sequences belonged to the same genetic cluster fell from 36.6% (95%CI: 30.6 - 45.1) when they were found within <100 km of each other to 5.7% (95%CI: 2.5 - 9.7) when they were 500-1,000 km apart. Using logistic regression, we found that each additional 100 km in spatial distance separating roosts was associated with 0.75 (95%CI: 0.73-0.77) times the odds of being part of the same genetic cluster (Figure 3E). As the probability of being from the same bat species is strongly linked to the distance between locations, we could not disentangle the role of spatial segregation between lineages from being solely a spatial effect or due to the different *Pteropus* species occupying different locations. However, on average, we found that pairs of sequences coming from the same bat species had 19.3 (95%CI 15.9 - 23.6) times the odds of belonging to the same genetic cluster (Figure 3F). Importantly, this characterisation of the changing probability of being from the same genetic cluster as a function of distance is robust to biased observation processes, and therefore can be considered a spatial signature of NiV ecology. Our estimates of spatial dependence were robust to broad variation in the definition of a genetic cluster that results in greater or fewer clusters (Figure S5).

To estimate the average number of genetic clusters circulating within an area, we used Approximate Bayesian Computation to fit our observed distribution of spatial clustering of genetic clusters (Figure 4A). We made a simplifying assumption that the spatial footprint covered by a genetic cluster is equivalent throughout the region, and that it can be captured by a multivariable normal distribution. We estimate that, on average, 95% of the infections from a genetic cluster were found within an area of 1.3 million km^2^ (95%CI: 0.6-2.3) and that there were an average of 14 discrete genetic clusters per land area of 100,000 km^2^ (95%CI: 6 - 24). We explore different ways to approximate the circulation area of *Pteropus* bats. For each country in the region, we estimate NiV diversity using existing estimates of *Pteropus* bats’ geographic range (from the International Union for Conservation of Nature [IUCN]). We also consider the area covered by tree cover (available from Global Forest Watch) as an alternative, indirect marker of the area covered by *Pteropus* bats. For this second approach, we separately consider over 10%, 30%, or 50% of tree cover as a proxy of the area covered by *Pteropus* bats (Figure 4B, Table 1, Figure S4, Table S6) (*18–20*). For most countries, we found little variation among estimates of diversity using these different approaches. For example, in Thailand, we estimate 15 unique genetic clusters (95%CI: 6 - 26) using the IUCN’s geographic range, and 17 unique genetic clusters (95%CI: 7 - 29) using the area with over 10% of tree cover. In Bangladesh, these same options suggest the presence of 15 genetic clusters (95%CI: 7 - 29), or of 11 genetic clusters (95%CI: 4 - 20), respectively. Taking South and Southeast Asia as a whole, we estimate between 66 and 118 separate NiV genetic clusters using the different approaches (range of confidence intervals of 28-211), compared to 15 currently detected, suggesting ∼80-90% of circulating genetic clusters remain undetected.

**Figure 4:**
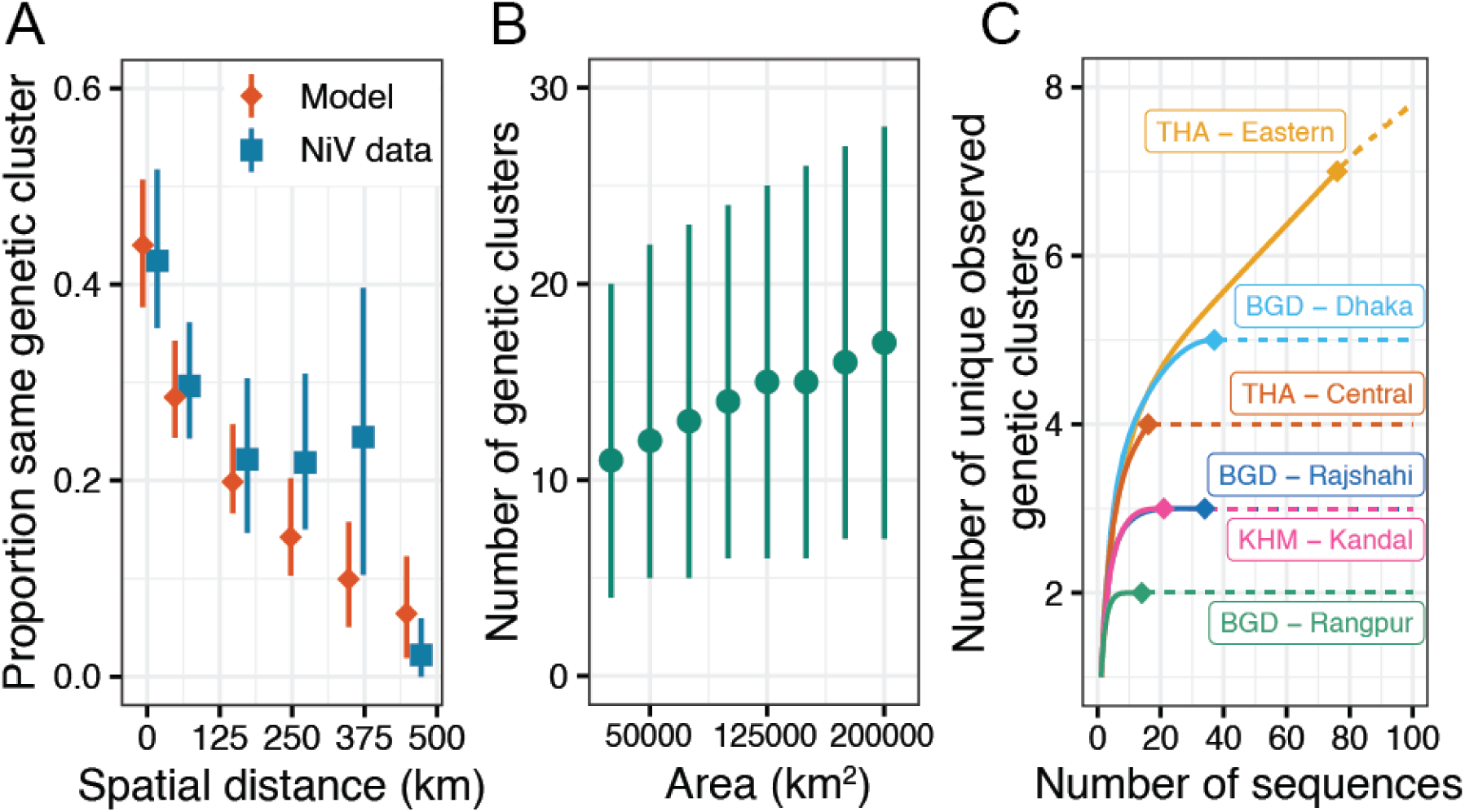
(A) Model fit using Approximate Bayesian Computation (ABC) on the proportion of sequence pairs that belong to the same genetic cluster in function of their spatial distance. The orange dots represent the median and the lines represent the 95% confidence intervals. Median proportions calculated on NiV data and used to calibrate the model are represented in blue. **(B)** Predicted number of genetic clusters in function of area (in square kilometers). The points and bars represent estimates of diversity for specific areas with 95% confidence intervals. **(C)** Estimated number of clusters in function of number of sampled NiV sequences for six regions in South and Southeast Asia. The dots represent the current number of observed samples and clusters in each region, the solid lines represent interpolated values based on observed data, and the dashed lines represent extrapolated values using Hill numbers of order 0.

**Table 1:** Estimations of the number of genetic clusters (with 95% Confidence Intervals in parentheses) for the six countries represented in our dataset, according to different estimates of *Pteropus* range (in km^2^): the IUCN’s estimations of *Pteropus* geographic range, and the Global Forest Watch’s estimations of tree cover per country above thresholds of 10%, 30%, and 50%, respectively.

|  | IUCN *Pteropus* range |  | Tree cover (10% threshold) |  | Tree cover (30% threshold) |  | Tree cover (50% threshold) |  |
| --- | --- | --- | --- | --- | --- | --- | --- | --- |
| Country | Area | N. clusters | Area | N. clusters | Area | N. clusters | Area | N. clusters |
| India | 2,987,747 | 55 (24, 89) | 490,910 | 23 (10, 37) | 388,304 | 21 (9, 34) | 304,653 | 19 (8, 31) |
| Bangladesh | 127,346 | 15 (6, 25) | 26,607 | 11 (4, 20) | 19,390 | 11 (4, 20) | 15,229 | 10 (4, 19) |
| Thailand | 138,538 | 15 (6, 26) | 219,195 | 17 (7, 29) | 199,624 | 17 (7, 28) | 179,669 | 16 (7, 27) |
| Cambodia | 62,957 | 13 (5, 22) | 101,985 | 14 (6, 24) | 88,099 | 14 (6, 24) | 74,823 | 13 (5, 23) |
| Malaysia | 323,929 | 20 (8, 32) | 297,861 | 19 (8, 31) | 294,306 | 19 (8, 31) | 289,772 | 19 (8, 31) |
| Indonesia | 1,656,164 | 40 (17, 63) | 1,650,987 | 39 (17, 63) | 1,606,412 | 39 (17, 62) | 1,550,634 | 38 (17, 61) |

Finally, we consider the extent to which genetic diversity has been fully uncovered in the six long term established surveillance sites in the region (three in Bangladesh, two in Thailand, and one in Cambodia), and whether there exist predictors of the number of genetic clusters in any place. There have been between 14 and 76 sequences obtained in these surveillance sites, resulting in the detection to date of between 2 and 7 different genetic clusters (Figure 4C). We estimated the number of new lineages that would be detected with additional sampling. This approach assumes that all clusters are equally likely to be detected and that all clusters circulating in an area are present at approximately equal levels. It also assumes stability in the clusters circulating in a location over time. We estimated that all the circulating genetic clusters have been identified in five of the six locations (Figure 4C, Figure S6). This can be directly inferred from the saturation in the number of observed clusters in those locations. The estimated total number of clusters circulating within a subnational division was not significantly associated with the size of the study area within the province, or with human population density. There was some evidence of an increase in diversity with percentage of forest cover (p-value of 0.015) (Figure S7).

## Discussion

We have used NiV sequences, alongside information on location and host from multiple countries to generate a comprehensive characterisation of the underlying genetic diversity of a pathogen that poses a major risk to human health. We found that NiV is strongly spatially structured. Overall, viral movement across the region is slow, with limited genetic similarity in the viruses circulating in different countries. These findings are consistent with a previous analysis using data from Bangladesh only (*21*). The evolutionary time separating sampled viruses in the two extremes of the Nipah region (i.e., India to Malaysia) was over 140 years. These patterns suggest that the virus is largely endemically entrenched within individual communities, with greatest diversity observed in the middle of this region. Cambodia was the only country in our dataset with all four sublineages detected. It remains unclear the extent to which the spatially structured nature of bat species, mixing patterns of bats across roosts and pre-existing immunity contribute to these observations.

The transmission dynamics of NiV within bat populations remains poorly understood, including whether infection results in long term immunity. In our study, we have found substantial overlap in the spatial footprint of genetic clusters, to the extent that even individual bat roosts host more than one distinct genetic cluster at any time. *Pteropus* bat roost size is highly dependent on the species, typically hosting around hundreds to thousands of bats at a time (*13, 22–24*). Maintaining a sufficiently large susceptible population to sustain multiple independent transmission chains in populations of this size will require long durations of viral shedding, frequent reinfection or coinfection as suggested through the modeling of bat immune profiles (*25*). As movement between bat roosts is common, the wider population across multiple roosts may also facilitate the maintenance of multiple independent lineages. In support of a key role of roost population size in maintaining diversity, it is notable that the *P. lylei* roosts in Cambodia, which had the greatest diversity with the co-circulation of three NiV sublineages within each roost, have typically thousands of bats in each roost, many more than other locations (*13*).

The evolutionary separation between NiV lineages has previously been suggested as one possible explanation for the differences in the case fatality rate in Bangladesh (CFR of ∼70%) and Malaysia (CFR of ∼40%) (*2, 26, 27*). However, it remains difficult to disentangle differences in the virus from human behavior differences or differences in transmission routes. Spillover from bats into pig populations through the consumption of NiV infected fruit drove the outbreak in Malaysia whereas date palm sap consumption by humans appears key in Bangladesh (*28*). Viral loads, inoculation routes and sites in these two transmission modes are likely to be very different, which could affect subsequent probabilities of death. Animal models such as primate, Syrian Hamsters and ferrets have not been able to provide conclusive evidence of pathological or transmission differences between the two major clades. However, their relevance to human situation remains unclear (*29–31*). Here we found evidence of differences in spillover or disease risk within a lineage from a single setting. Genotype IB was found in three different years in bat roosts in an area of Bangladesh where human spillovers are frequently identified and where date palm sap consumption is common. However, no human cases were linked to this sublineage. It remains possible that underlying year-by-year variability in spillover risk, linked to temperature, may explain these findings, especially in the context when only a subset of human NiV cases are ever detected and have their viruses sequenced (*8, 10, 32*).

Using the correlation of genetic clusters over distance as a spatial signature of NiV, we were able to estimate the spatial footprint of individual genetic clusters and the number of circulating clusters. While these are crude estimates, they do provide a ballpark estimate of the expected number of NiV genetic clusters, which is especially useful in countries where little or no sampling has been conducted. We estimated that across the entire region there are ∼100 genetic clusters. This suggests the vast majority (∼80-90%) of genetic diversity remains undetected. In particular, India is likely to harbor significant diversity. *Pteropus* bats are found throughout the country and have been found to be positive in serology and PCR detection to NiV throughout (*3*). Increased sampling across South and Southeast Asia will help uncover additional lineages that have not yet been observed. Extending the sampling to new locations will expand the observed diversity with limited new diversity likely to be found from additional sampling in the established long-term sampling locations in Cambodia, Thailand, and Bangladesh. Long-term surveillance in the same locations remains a critical source of information to characterize long-term evolutionary dynamics of NiV within roosts. Whole-genome sequencing should be prioritized to maximize the signal that can be captured through established and additional surveillance.

Obtaining NiV sequences from bats is difficult. The most common method requires the collection of bat urine under roosts using plastic or tarpaulin sheets. The urine can then be tested for evidence of virus using PCR tests. However, this approach makes it impossible to link urine to an individual bat. It is also impossible to be sure that multiple positive samples don’t all come from the same bat.

Most viral isolates come from urine samples taken from individual bats, which makes them very rare. For this reason, most available NiV sequences from bats are short sequences from the PCR process. Despite their frequent short nature, we were still able to consistently place sequences in different clades. In particular, in Cambodia, where most sequences were very short (<400 nucleotides in length), we were able to identify multiple clades circulating within the same roosts. As with other phylogenetic studies, this study is reliant on the available sequences, which have been collected in a spatially and temporally biased manner. However, the geographic clustering of genetic clusters is robust to these biases. The classification of tips into genetic clusters necessarily relies on thresholds of evolutionary distance, however, in sensitivity analyses, changes in these thresholds resulted in minimal changes to the overall inferences on the genetic diversity of viruses circulating within any area. Finally, NiV sequences are ultimately reliant on the PCR primers used to detect the virus in the original sample. If the PCR primers are overly specific, they may systematically miss some viruses (*33*). Future efforts may want to consider using broader primer sets.

This project has demonstrated that even sparsely sampled genetic data, including many short sequences, from large areas can provide meaningful characterization of underlying diversity in populations when considered together. We have shown that even individual roosts typically have multiple circulating transmission chains, but with each genetic lineage covering a large spatial footprint, probably driven by bat mobility patterns. While most NiV diversity remains undetected, the surveillance sites which have been established appear to have uncovered a substantial proportion of the diversity in those locations.

## Materials and Methods

### Data collection and alignment

We collected all available NiV genomes in GenBank (n=301) (*34*) and compiled their date and place of collection, and the host species they were sampled from. We also included several previously unpublished sequences (n=26), collected between 2013 and 2016 in two bat roosts in Cambodia. The sampling and screening approach for NiV for these additional sequences is explained elsewhere (*13*).

Sequence alignment was performed using MUSCLE and manually corrected using MEGA-X (*35*). 70 sequences were excluded because of poor quality, because they were referenced on GenBank twice, or because they were different sequenced genes from one single sample (in which case they were combined into one genome). The resulting data set included 257 sequences from 6 countries (India, Bangladesh, Thailand, Cambodia, Indonesia, and Malaysia, see Figure 1A), spanning over a range of 22 years (1999–2020) (Table S1) (*5, 12, 21, 30, 36–55*). 175 sequences were sampled from six different bat host species: *Pteropus lylei* (n=120)*, Pteropus medius* (formerly *Pteropus giganteus*, n=41)*, Pteropus vampyrus* (n=6)*, Pteropus hypomelanus* (n=6), *Hipposideros larvatus* (n=1), and one sequence came from a bat in the *Taphozous* genus, though its exact species is undetermined (Figure 1B) (*55*). The rest of the sequences were sampled from humans (n=73), pigs (n=7), and one sequence from a dog. 1 sequence had an uncertain host. Sequence length varies from 153 bp to 18.2k bp (full genome). In total, there are 64 full-length genomes, 185 genomes covering (at least partially) the nucleocapsid (N) gene, and the rest cover different parts of the NiV genome. Sequence lengths are detailed in Supplementary Table S2.

### Preliminary Analyses

We ran a preliminary analysis using IQ-TREE (v 1.6.12) (*56*) in order to select the best-fitting nucleotide substitution model. The model was selected through a bootstrap with 1000 resampling events. According to the Bayesian Information Criterion (BIC), the TIM2 model was selected. We performed Bayesian Evaluation of Temporal Signal (BETS) (*57, 58*) to determine if the temporal resolution of our NiV alignment was sufficient to calibrate a molecular clock. We compared two models: one in which the sequences were considered heterochronous, using their real sampling dates, and another one in which the sequences were considered isochronous, with a fixed evolutionary rate. The marginal log-likelihoods for both models were estimated using stepping-stone sampling (*57*), and they were compared to obtain a Bayes factor.

### Phylogenetic reconstruction

We reconstructed a time-resolved phylogeny using BEAST v.1.10.4 (*59*), which implements MCMC to analyze time-explicit genetic sequences. We used a subset of 172 sequences, comprising only unique sequences per year and country. For each sequence, we specified the sampling date in the decimal year format, using the most precise information available (in GenBank or the corresponding publication). For sequences that we only knew the year of sampling (n=29), we added a one-year uncertainty around the specified time. We implemented three parallel runs of six consecutive chains of 200M states each. We used the generalized time reversible substitution model (GTR), as it was the only evolutionary model proposed by BEAST that offered at least as many degrees of freedom as the TIM2 model. We used a lognormal uncorrelated relaxed molecular clock, and the Bayesian Skygrid (*60*) coalescent tree prior. The chains were sampled every 20,000 states. For each of the three parallel runs, the six consecutive chains were combined into one and resampled with a frequency of 320,000 states using the LogCombiner software (*61*) from the BEAST suite.

We visualized the posterior distribution using Tracer (*62*), and obtained satisfactory ESS values (>200) for all estimated parameters. Based on the tree files combined in the same way as described in the previous paragraph, we summarized a maximum clade credibility (MCC) tree using TreeAnnotator (*63*). We visualized the MCC tree using the FigTree software (*64*) and the ggtree package in R v.4.2.0 (*65–69*).

### Initial phylogenetic classification

Based on the reconstructed phylogeny’s structure, and on previous phylogenetic analyses of NiV genomes (*15*), we classified the NiV genomes into two major lineages (lineages I and II, corresponding to the lineages that have been previously described as the Bangladeshi and the Malaysian lineages, respectively), and four sublineages (I.A, I.B, II.A, II.B, see Figure 1C). Using this broad genetic classification, we analyzed NiV’s spatial structure across South and Southeast Asia by looking at the sublineage distribution per country, organizing countries according to their longitude (Figure 1D).

### Distribution of human cases in Bangladesh

The distribution of human sequences in the reconstructed phylogeny is of interest to estimate if there are sublineages that are more prone to spillover events. We investigated the distribution of human sequences coming from Bangladesh and compared it to the distribution of bat sequences (Figure S2), because this is the only country where human behavior results in regularly documented spillovers (*2*). For sequences coming from Bangladesh, we considered only one human sequence per case cluster, and only one bat sequence per district-year. Sequences sampled from humans collected in 2004 were considered as part of the same cluster if they were sampled in the same district and the same year, otherwise they were considered from different case clusters. For human sequences sampled in 2008 and 2010, outbreak information was extracted from (*5*). For human sequences sampled in 2011, information about case clusters was extracted from (*70*). For human sequences sampled from 2012 onwards and included in (*12*) (n=19), the publication indicates if the human case was primary or not, if it was clustered with other cases and, in most cases, which other sequences belonged to the same case cluster. Two sequences from 2014 that were marked as clustered, with no specification about whether they belonged to the same cluster or not, and that were sampled in the same subdistrict on the same day, were considered as part of the same case cluster and only one of them was randomly selected and included at this stage of the analysis. The rest of human sequences sampled after 2012 either did not have any precise location information, in which case they were excluded from this part of the analysis (n=3), or they were published in (*21*) (n=14), and they were treated in the same way as 2004 sequences.

This resulted in a dataset of 38 sequences, with 30 human sequences, all of which came from sublineage I.A, and 8 bat sequences, out of which 5 (62.5%) came from sublineage I.A and 3 (37.5%) came from sublineage I.B. The purpose of this data subsetting is to avoid multiple sequences that are identical or too closely related from one single outbreak or roost visit. Due to the small size of this subset, we implemented Fisher’s exact test to analyze the significance of the contingency between human cases and each of the aforementioned sublineages.

### Spatial spread

In order to analyze the speed at which NiV has spread across South and Southeast Asia, we computed the mean spatial pairwise distance as a function of the pairwise evolutionary distance for NiV sequences. We used a subset in which, for sequences sampled from bats, we only included one sequence per roost, per year (n=44). For sequences sampled from human infections, we only included one sequence per outbreak (n=34). This resulted in a subset of 78 sequences. For each sequence, we used the coordinates of the most precise sampling location available. For bat sequences, this was either the bat roost location (n=32), or the centroid of the district where the sequences were collected (n=12). We computed the geodesic pairwise spatial distances using the geodist package in R (*71*). We computed the pairwise evolutionary distance for all pairs of sequences using the cophenetic.phylo function from the ape package on the reconstructed phylogeny (*72*). We then computed the mean pairwise spatial distance for pairs of sequences separated by different cumulative evolutionary distances (Figure 2), implementing a non-parametric bootstrap with 1000 resampling events.

### Phylogenetic clustering using a finer genetic resolution

We used the PhyCLIP module in Python to cluster the sequences in the tree into different lineages (Figure 3D) (*16*). PhyCLIP aims to do phylogenetic clustering based on statistically informed rather than user-defined divergence thresholds, with a linear integer optimization approach. The module takes a phylogeny as input, along with three parameters:

- *S*, the minimum number of sequences that can constitute a cluster,
- *γ*, the multiple of deviations from the grand median of the mean pairwise distance that defines the within-cluster divergence limit, and
- *FDR*, a false discovery rate to ensure that every pair of clusters are significantly different from one another.

We ran PhyCLIP using a combination of different values for each of these parameters (S ∈ {1,2,3}, *γ* ∈ {1, 2, 3}, *FDR* ∈ {0.1, 0.15, 0.2, 0.25, 0.3}), using the module’s optimisation criteria to find an optimal combination of parameter values. With the aim of doing a comprehensive clustering of all available NiV sequences, we chose PhyCLIP’s intermediate optimization resolution (as opposed to high resolution), which optimizes the values of the parameters based on the maximum percentage of sequences assigned to a cluster, the minimum grand mean of within-cluster divergence, and the maximum mean inter-cluster divergence. We compared the clustering results of all sets of parameters that resulted in more than 90% sequences belonging to a cluster to the optimal solution determined by the criteria above using the Adjusted Rand Index from the fossil package in R ((*17*) (*73*)). We repeated this on 100 randomly sampled trees from our BEAST posterior, comparing the top results to the optimal solution on our MCC Tree.

Then, for sets of parameters that resulted in more than 90% sequences belonging to a cluster (Supplementary Table S3), we manually corrected PhyCLIP’s output to assign a cluster to all outliers, and to ensure that all clusters were monophyletic. Finally, we compared the resulting options for the classification of the NiV phylogeny, by computing the proportion of pairs of sequences belonging to the same cluster in function of their spatial distance (Supplementary Figure S5). Since there was little variation between these different options, we selected the classification into 15 clusters, as this corresponded to PhyCLIP’s optimal solution (Figure 3D).

### NiV cluster distribution

We analyzed the spatial and host species structure for sequences sampled from bats (n=175) using this finer phylogenetic classification (Figures 3A-C). Duplicate sequences that were not included in the BEAST or the PhyCLIP analyses were assigned the same cluster as the sequence that they were identical to from their country and year. For each sequence cluster, we analyzed its spatial distribution at a country scale (Figure 3A), and its distribution across all bat host species (Figure 3B). We then analyzed the relationship between these by looking, for each country, at the bat species distribution in our dataset (Figure 3C).

### Models of cluster correlation

We developed a set of models to investigate the contribution of different covariates to the probability that a pair of sequences belong to the same genetic cluster. For pairs of sequences, we analyzed the effects of coming from the same location, of distance, and of coming from the same host species or not. Due to high correlation between the spatial and the host species covariates, we chose to separate them into two distinct models. We show the relationship between these two covariates by fitting a logistic model of the probability that a pair of sequences comes from the same host species to their spatial distance (Figure S3).

### Spatial model

For all pairs of bat sequences with roost (n=132) or district information (n=36), we considered:

- Whether the sequences came from the same location or not (*l*),
- For sequences coming from different locations, we also considered their spatial distance (*d*),

As shown in Equation 1:

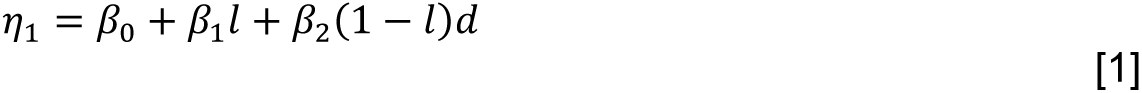

We then used these parameters to fit a logistic regression to estimate the probability P that a pair of sequences comes from the same genetic cluster (Equation 2, see Figure 3E):

### Host species model

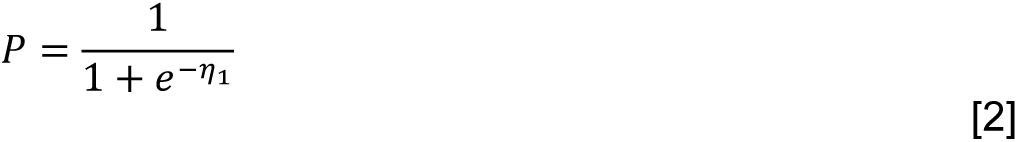

Separately, for all pairs of bat sequences, we fit a logistic model of the probability that a pair of sequences comes from the same genetic cluster depending on whether the sequences came from the same host species or not (Figure 3F).

### Assessment and prediction of diversity

We investigated the way in which the observation of different genetic clusters evolved with sampling in specific areas of South and Southeast Asia. We focused on divisions with at least 10 sequences, and in which at least 2 clusters have been observed: Dhaka, Rajshahi, and Rangpur divisions in Bangladesh, Kandal in Cambodia, and Eastern and Central Thailand. We used the iNEXT package (*74, 75*), with which we implemented a rarefaction analysis using Hill numbers of order 0 (equivalent to Species Diversity) to interpolate how many clusters had been observed in each division since sampling began until the current number of observations in each place, and to extrapolate how many more we could expect to observe if more NiV sequences were sampled in these regions. Average estimates can be seen on Figure 4C and estimates with 95% Confidence Intervals can be seen on Supplementary Figure S6.

We investigated the relationship between the estimated number of clusters in these six regions and several ecological variables, with the aim of exploring if any of these covariates could be used to provide an estimation of the number of NiV genetic clusters in regions where sampling was not sufficient to conduct a rarefaction analysis. We looked at the mean pairwise geographic distance between sequences, at the human population density for each region (*76–78*), and at the percentage of tree coverage for each region (Supplementary Table S4, Supplementary Figure S7). For the tree coverage estimation, we averaged the estimates of coverage over a threshold of 30% for 2000 and 2010 from the Global Forest Watch (*19, 20*). We explored these measures as potential proxies for bat mobility, for human pressure on bat roosts, and for bat population size in each of these provinces, respectively. We plotted the estimated number of clusters for each of the six regions as a function of these parameters, and we fit a generalized Poisson regression for each of them.

### Prediction of NiV diversity using Approximate Bayesian Computation

Approximate Bayesian Computation (ABC) is a method used when no explicit likelihood function can be written for a model, or when such a function would be too costly in terms of time or computational resources (*79*). Considering the amount of underlying unknowns in terms of bat mobility and of interactions between different bat roosts and bat species, we adopted this approach to further investigate ways in which the spatial footprint of NiV genetic clusters can be captured with the aim of providing a rough estimate of how many different genetic clusters can be found in a specific area.

We computed a set of summary statistics on the NiV data set. For a specific spatial window wi, we compute the median proportion i of sequence pairs, within that window, that belong to the same cluster:

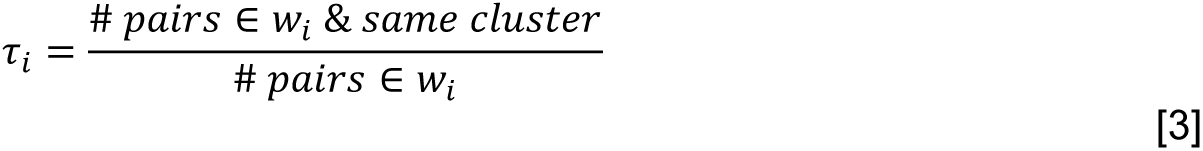

We developed a simulation framework for the spatial footprint of NiV genetic clusters in a specific area. The input for a simulation was the cluster density *clust*_*dens*_(in clusters·km^-2^), and the standard deviation *sd*_*clust*_, a measure of the spread of each genetic cluster. In each simulation:

- We generated a polygon (square) as our area of study, of width *widt*ℎ_*outer*_.
- Using the size of the polygon and the cluster density *clust*_*dens*_, we computed the number of clusters to simulate.
- We randomly sampled the x and y coordinates of each cluster “center” within the initial polygon using a uniform distribution (Figure S8A).
- For each cluster, we randomly sampled *n*_*obs*_ points. The distance from each point to its corresponding cluster center was sampled using a truncated normal distribution of mean 0 and of standard deviation *sd*_*clust*_, with a truncation at two times the standard deviation, allowing for 95% of the normal distribution to be sampled. The angle was sampled using a uniform distribution between 0 and 2*π* (Figure S8B).
- Then, we subset these simulated points to keep only those points within a sampling square of width *widt*ℎ_*inner*_ and centered inside the initial square (Figure S8C).
- Finally, we computed the summary statistics *τ*_*sim*,*i*_ for each spatial window in the same way as with the NiV dataset.

We implemented this framework using Python 3.8 (*80*). We optimized the computation of the summary statistics for each simulation by partitioning the sampling square into a grid. Thus, with a grid width greater than the maximum distance for which *τ*_*sim*,*i*_ is computed, points within a specific cell only needed to be compared to points within their own or neighboring cells (see Figure S8D).

Distance computation was performed using a Nearest Neighbors algorithm from the scikit-learn library v.1.2.1 (*81*). Parameter prior distributions and hyperparameter values are detailed in Table S5.

We computed the residual sum of squares for each of the *n*_*sim*_ simulations, weighting each spatial window by the number of pairs within it in our actual NiV dataset. We then kept the 2% closest simulations. Model fit can be seen in Figure 4A.

We used this final set of simulations to compute the average number of unique observed clusters per area. For each simulation, and for each specific area, we randomly placed 1000 circles of the corresponding area in the simulated space, we counted the number of clusters that were contained within or that intersected each circle, and we computed the average of observed clusters per area across all simulations (Figure 4B).

Finally, we used this way of estimating diversity in function of area to compute rough estimates of the number of clusters that can be observed in the countries represented in our dataset. We explored different options for the area of circulation of *Pteropus* bats. We downloaded the geographic range estimates from the International Union for Conservation of Nature’s Red List of Threatened Species (IUCN) (*18*), and we computed the coverage in km^2^ of *Pteropus* bats per country, for all countries with *Pteropus* bat circulation. We also used an estimation of tree cover area for each country, since this represents the ecosystem where *Pteropus* bats live (*8*). For each country, we computed the mean tree cover in km^2^ over a threshold of 10%, 30%, and 50% for 2000 and 2010 from the Global Forest Watch (Table S6, Figure S8) (*19, 20*).

## Data Availability

Most data referred to in the manuscript are available online on GenBank (accession numbers are provided in Supplementary Table S1). This study includes some previously unpublished sequences, which will be made available on GenBank upon acceptance of the manuscript.

https://www.ncbi.nlm.nih.gov/nuccore/?term=nipah+henipavirus

## Supplementary Information

**Figure S1:**
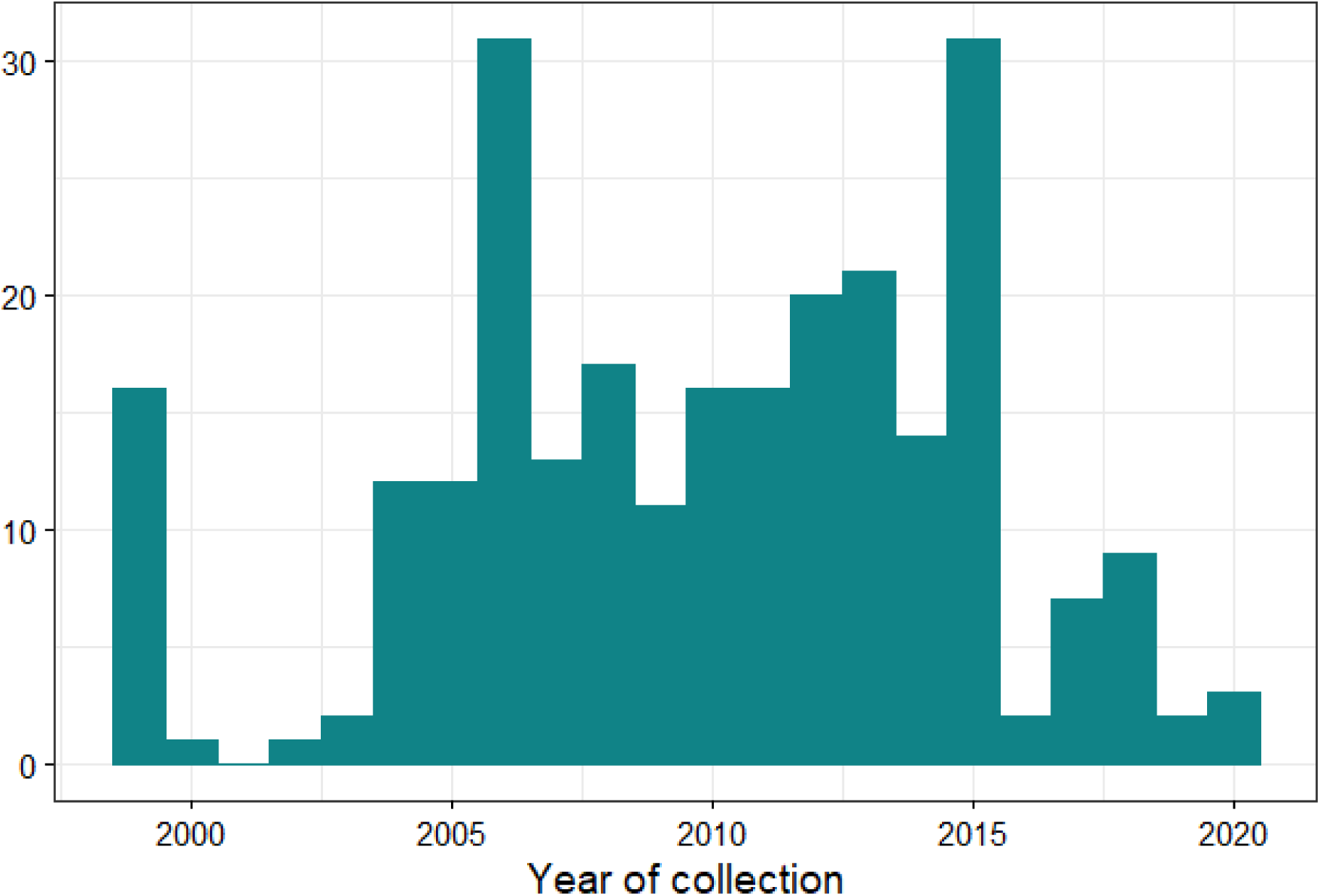
Histogram of the years of collection of sequences.

**Figure S2:**
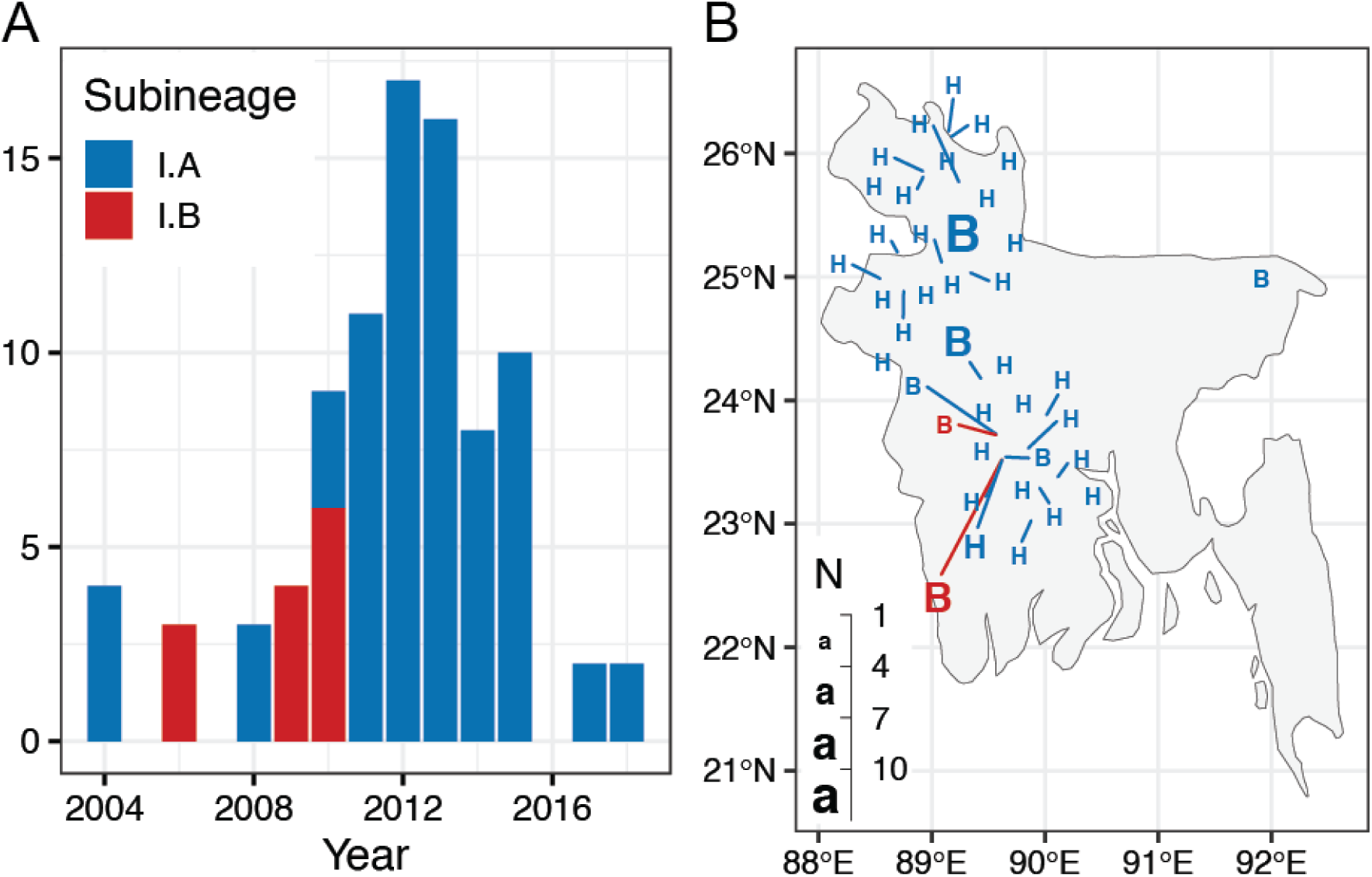
(A) Temporal distribution of Bangladeshi sequences, colored according to their sublineage. **(B)** Spatial distribution of Bangladeshi sequences. “H”s represent sequences sampled from humans, and “B”s represent sequences sampled from bats.

**Figure S3:**
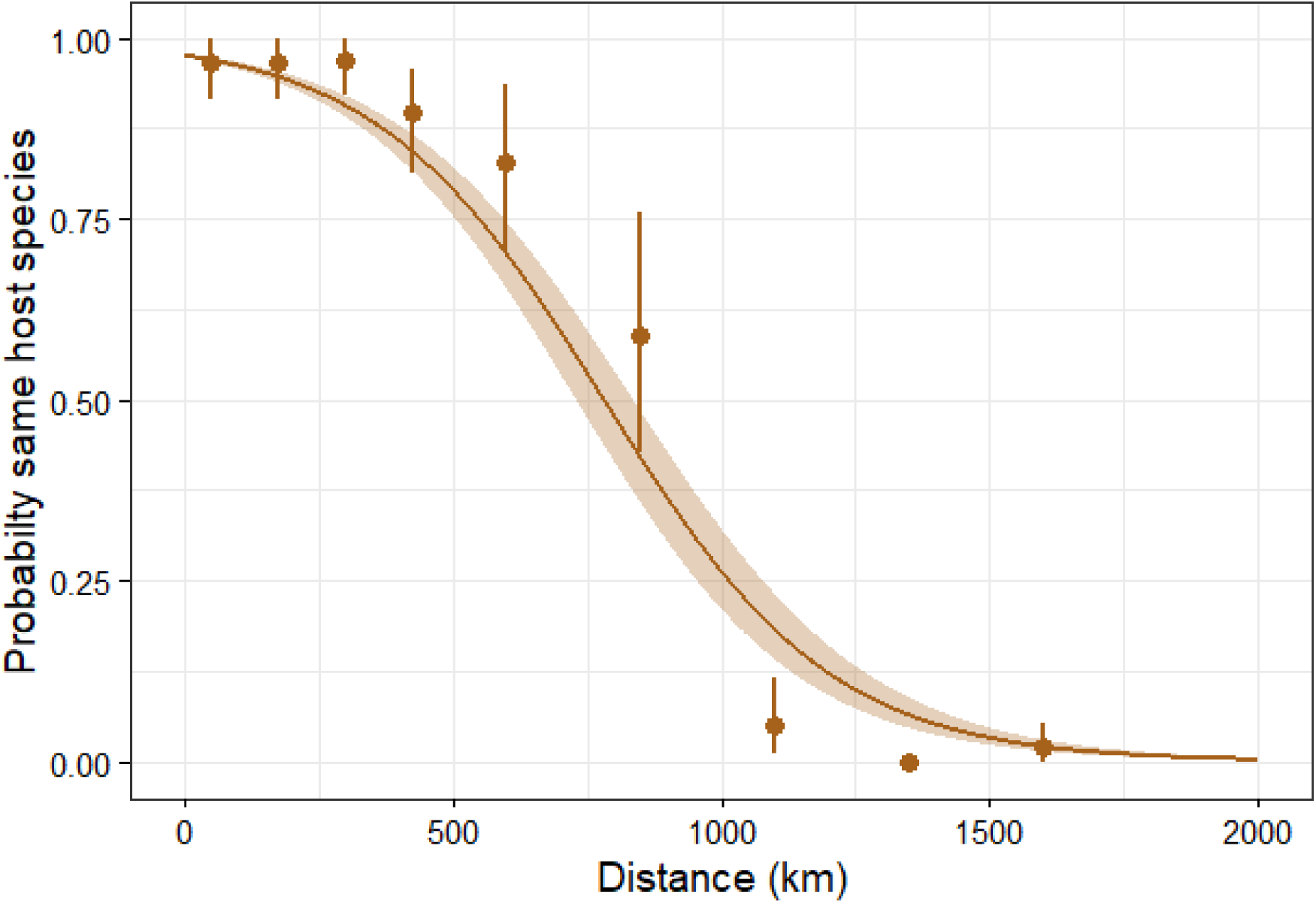
Proportion of sequence pairs coming from the same host species as a function of their spatial distance, with a logistic fit. The brown shaded region represents 95% confidence intervals.

**Figure S4:**
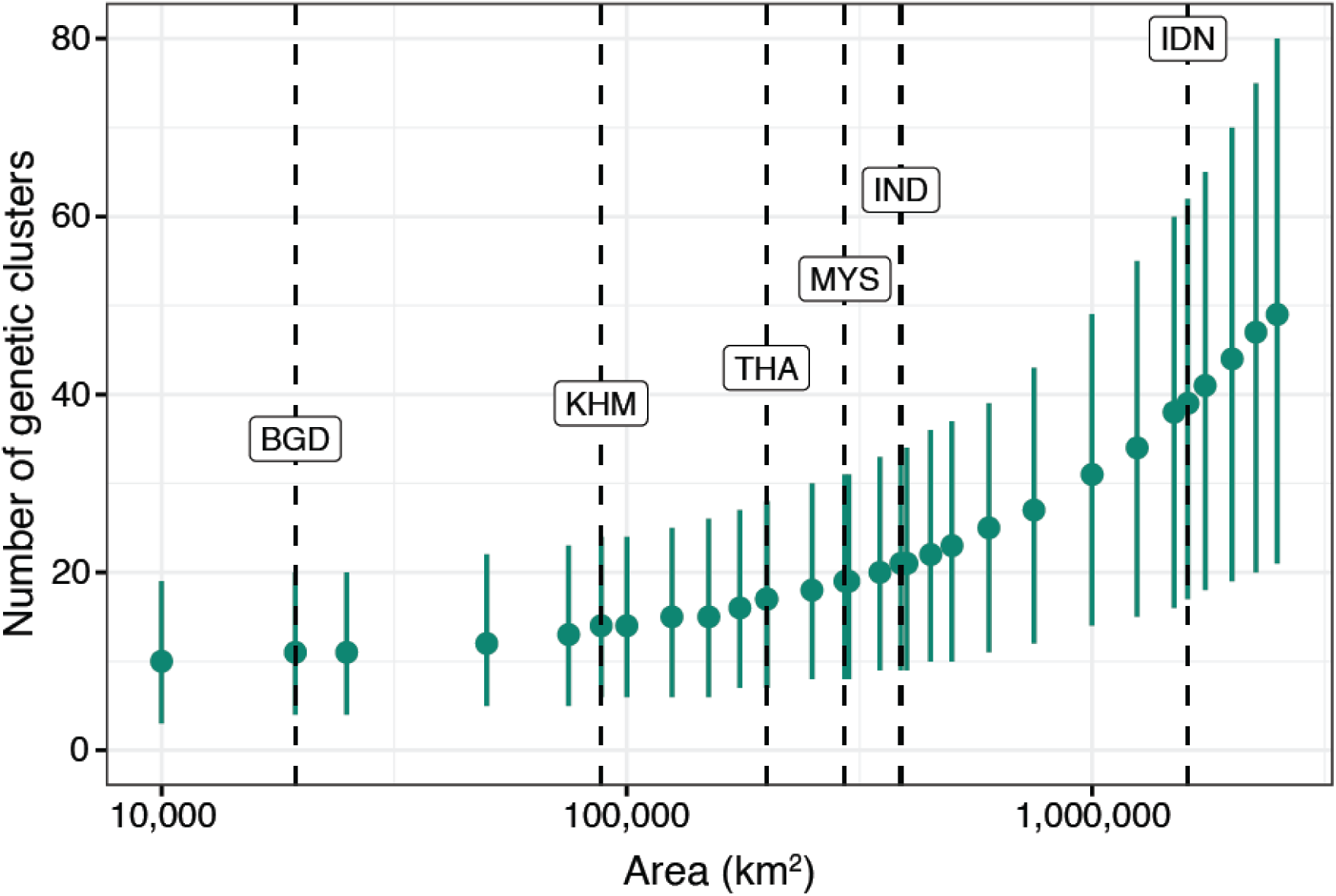
Estimates of numbers of NiV genetic clusters per area. Dashed lines represent the estimates for countries represented in our dataset - Bangladesh, Cambodia, Thailand, Malaysia, India, and Indonesia - based on each country’s tree-covered areas.

**Figure S5:**
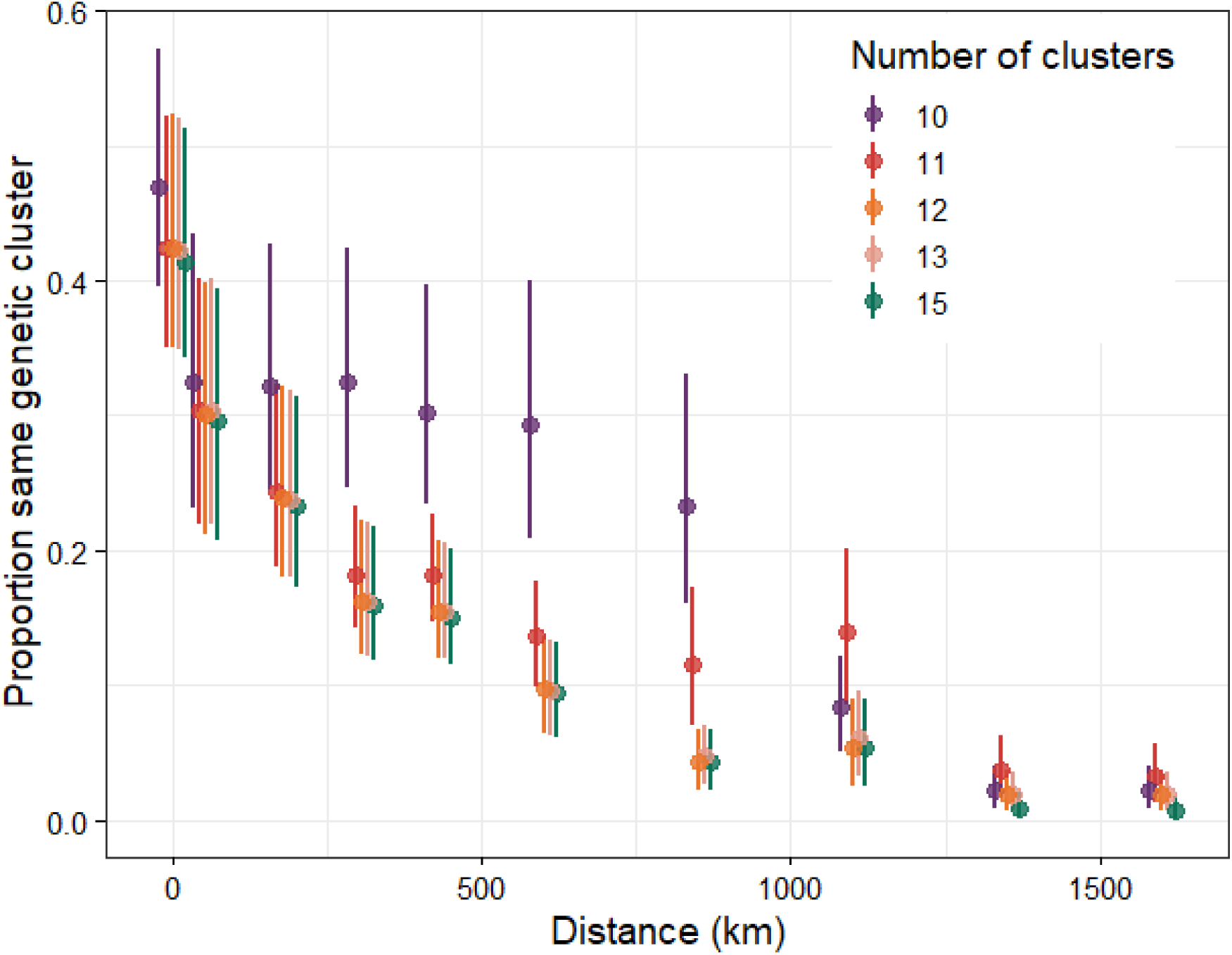
Sensitivity analysis on the pairwise probability that two sequences belong two the same genetic cluster in function of the spatial distance separating them, computed for different clustering options of the NiV phylogeny.

**Figure S6:**
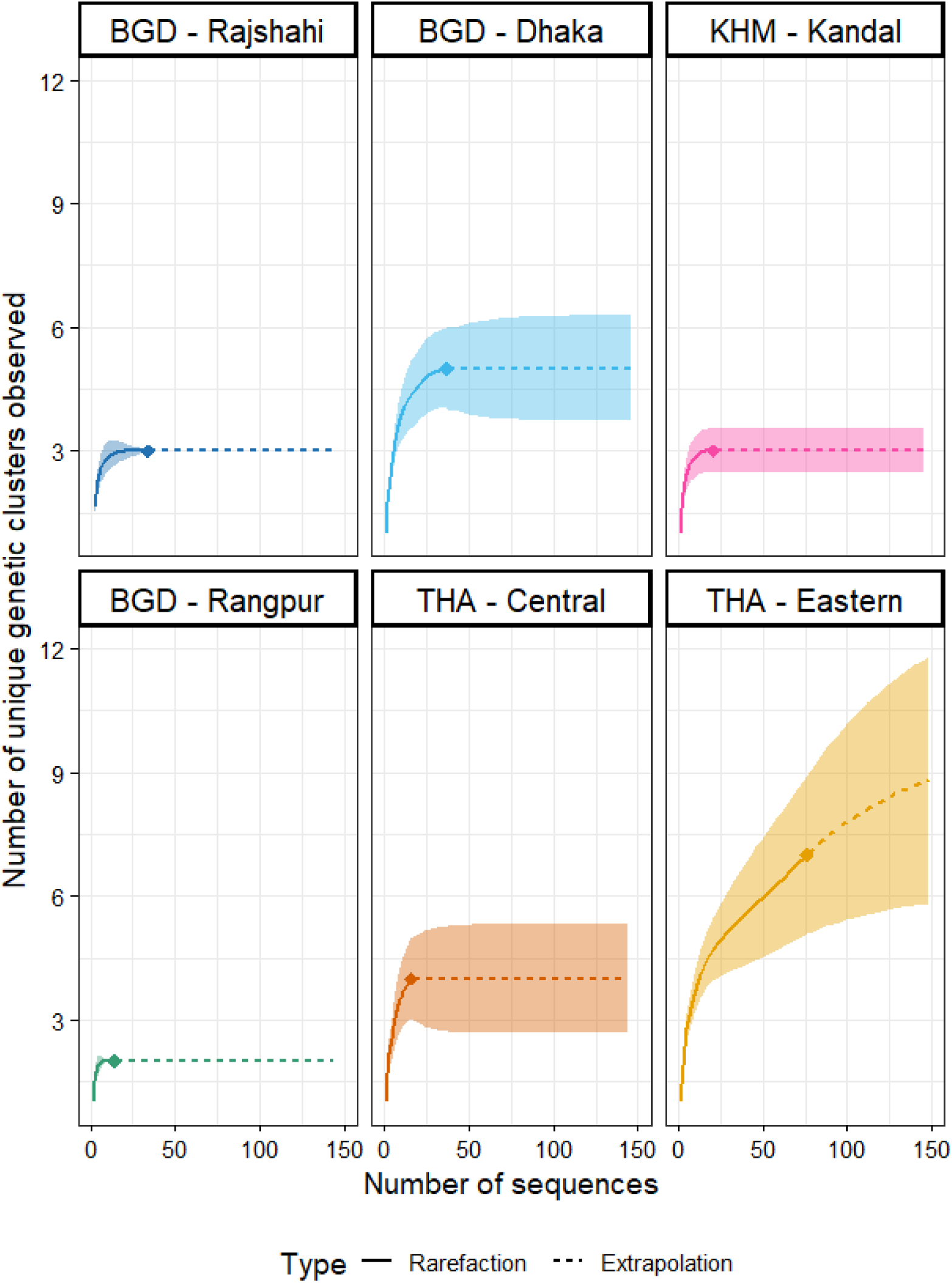
Estimated number of clusters in function of number of sampled NiV sequences for six regions in South and Southeast Asia. The points represent the current number of observed samples and clusters in each region, the solid lines represent interpolated values based on observed data, and the dashed lines represent extrapolated values using Hill numbers of order 0 using the iNEXT package on R. Ribbons represent 95% Confidence Intervals.

**Figure S7:**
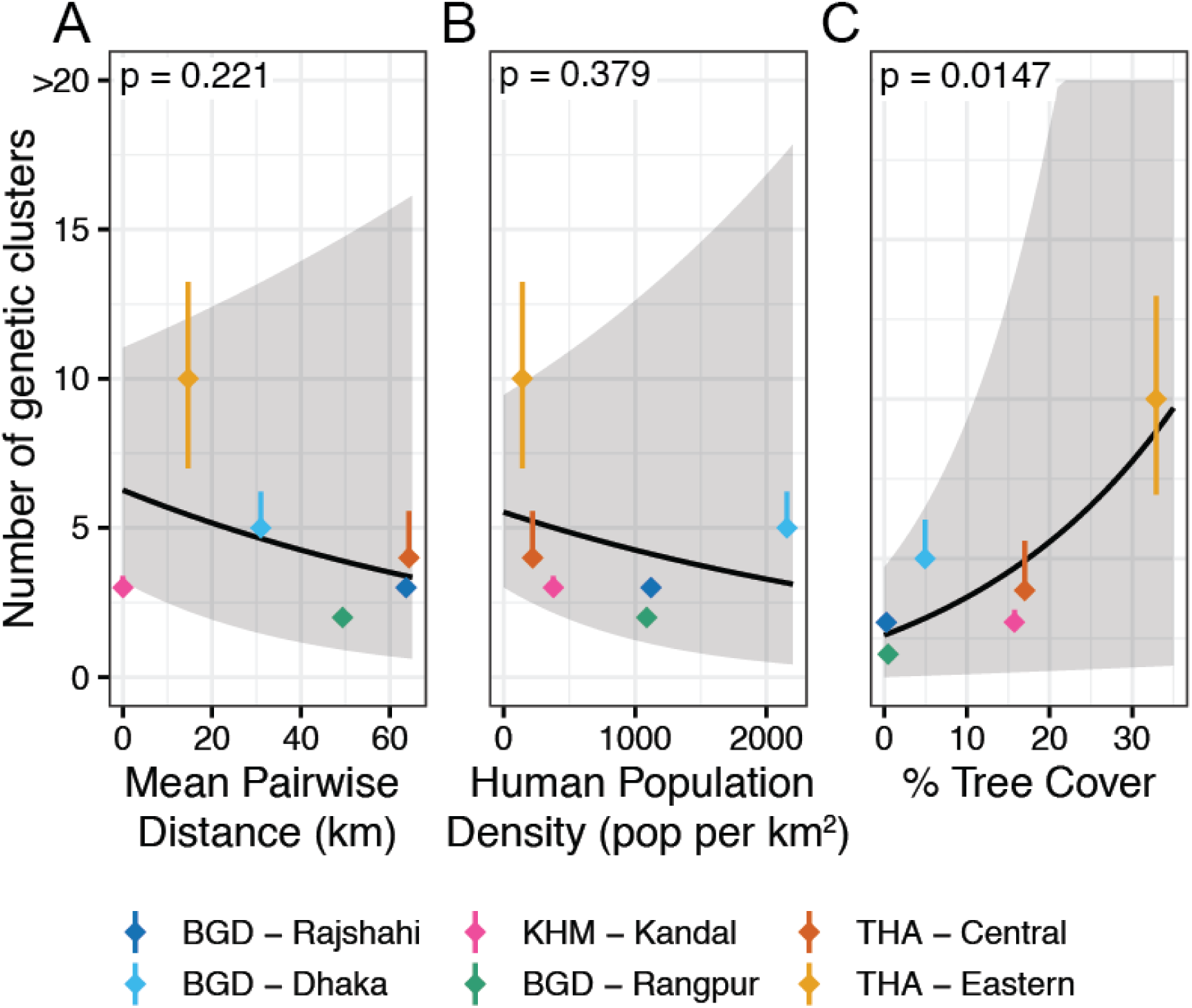
Estimated asymptotic number of genetic clusters for six regions in South and Southeast Asia in function of: **(A)** the mean pairwise geographic distance separating sequences sampled from these regions, **(B)** the regional human population density (in inhabitants per km^2^), and **(C)** the regional tree cover percentage, with a Poisson Generalized Linear Model fit.

**Figure S8:**
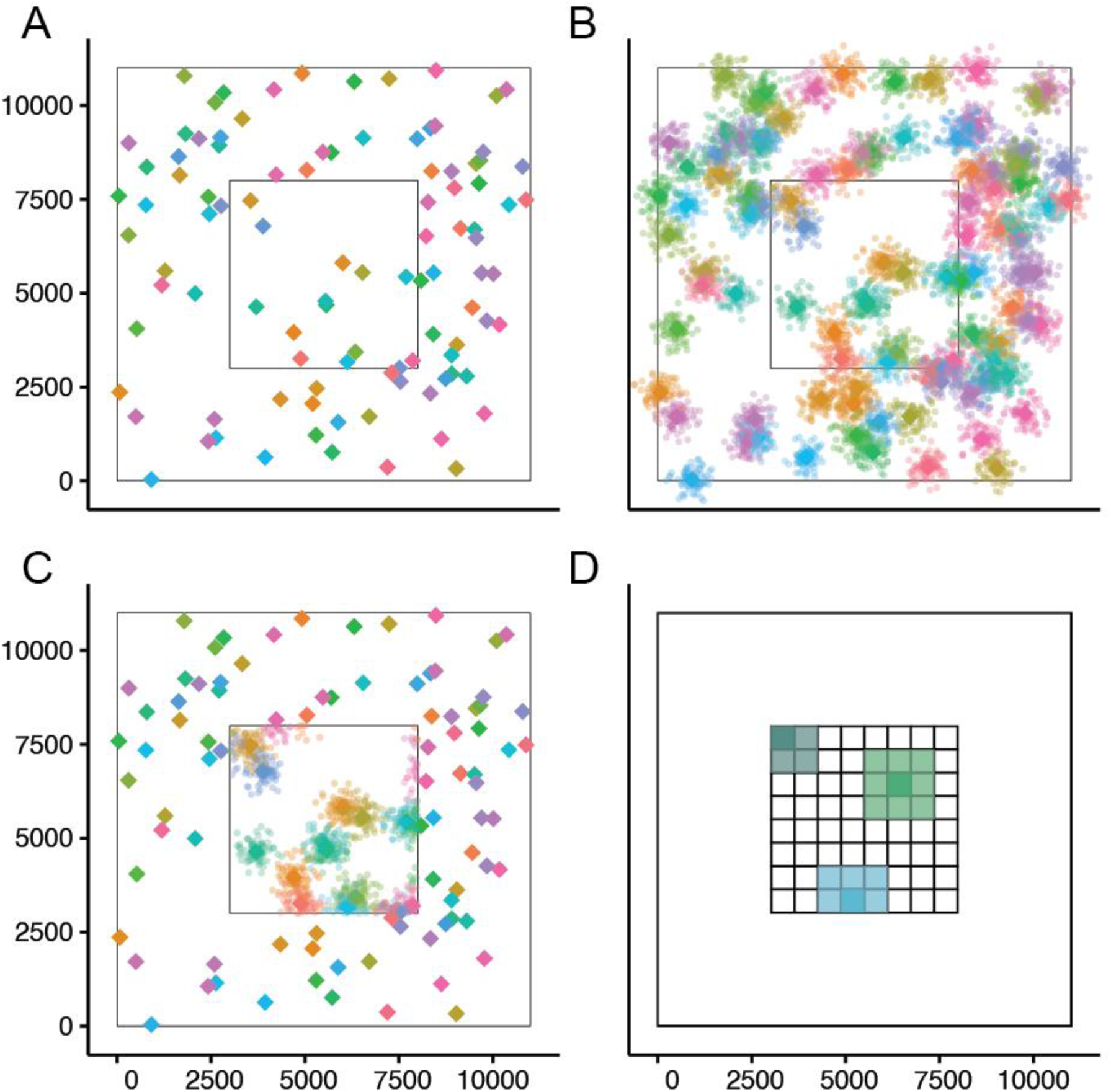
(A) Simulated centres of genetic clusters, **(B)** simulated samples from each cluster using a truncated normal distribution, **(C)** simulation of sampling by subsetting samples within a secondary polygon, **(D)** spatial partitioning of sampling square into a grid to optimize the computation time of summary statistics for each simulation.

**Table S1:** GenBank Accession number, country of provenance, and host species for all sequences used in this study (n=257).

| **Accession** | **Country** | **Host Species** |
| --- | --- | --- |
| NC_002728 | Malaysia | *Unspecified* |
| KY887671 | Bangladesh | *Homo sapiens* |
| KY887670 | Bangladesh | *Homo sapiens* |
| KY425655 | Malaysia | *Homo sapiens* |
| KY425646 | Malaysia | *Homo sapiens* |
| FJ648082 | Thailand | *Pteropus lylei* |
| FJ648081 | Thailand | *Pteropus lylei* |
| FJ648080 | Thailand | *Pteropus lylei* |
| FJ648079 | Thailand | *Pteropus lylei* |
| FJ648078 | Thailand | *Pteropus lylei* |
| FJ648077 | Thailand | *Pteropus lylei* |
| FJ648076 | Thailand | *Pteropus lylei* |
| FJ648075 | Thailand | *Pteropus lylei* |
| FJ513081 | India | *Homo sapiens* |
| FJ513080 | India | *Homo sapiens* |
| FJ513079 | India | *Homo sapiens* |
| DQ061858 | Thailand | *Pteropus lylei* |
| DQ061857 | Thailand | *Pteropus lylei* |
| DQ061856 | Thailand | *Pteropus lylei* |
| DQ061855 | Thailand | *Pteropus lylei* |
| DQ061854 | Thailand | *Pteropus lylei* |
| DQ061853 | Thailand | *Pteropus lylei* |
| DQ061852 | Thailand | *Pteropus lylei* |
| DQ061851 | Thailand | *Hipposideros larvatus* |
| EF070190 | Thailand | *Pteropus lylei* |
| KT163257 | Thailand | *Pteropus hypomelanus* |
| KT163256 | Thailand | *Pteropus lylei* |
| KT163255 | Thailand | *Pteropus lylei* |
| KT163254 | Thailand | *Pteropus lylei* |
| KT163253 | Thailand | *Pteropus lylei* |
| KT163252 | Thailand | *Pteropus lylei* |
| KT163251 | Thailand | *Pteropus lylei* |
| KT163250 | Thailand | *Pteropus hypomelanus* |
| KT163249 | Thailand | *Pteropus hypomelanus* |
| KT163248 | Thailand | *Pteropus hypomelanus* |
| KT163247 | Thailand | *Pteropus hypomelanus* |
| KC903172 | Indonesia | *Pteropus vampyrus* |
| KC903171 | Indonesia | *Pteropus vampyrus* |
| KC903170 | Indonesia | *Pteropus vampyrus* |
| KC903169 | Indonesia | *Pteropus vampyrus* |
| KC903168 | Indonesia | *Pteropus vampyrus* |
| JF899339 | India | *Pteropus medius* |
| EU624737 | Thailand | *Pteropus lylei* |
| EU624736 | Thailand | *Pteropus lylei* |
| EU624735 | Thailand | *Pteropus lylei* |
| EU620498 | Thailand | *Pteropus lylei* |
| EU603758 | Thailand | *Pteropus lylei* |
| EU603757 | Thailand | *Pteropus lylei* |
| EU603756 | Thailand | *Pteropus lylei* |
| EU603755 | Thailand | *Pteropus lylei* |
| EU603754 | Thailand | *Pteropus lylei* |
| EU603753 | Thailand | *Pteropus lylei* |
| EU603752 | Thailand | *Taphozous* |
| EU603751 | Thailand | *Pteropus lylei* |
| EU603750 | Thailand | *Pteropus lylei* |
| EU603749 | Thailand | *Pteropus lylei* |
| EU603748 | Thailand | *Pteropus lylei* |
| EU603747 | Thailand | *Pteropus lylei* |
| EU603746 | Thailand | *Pteropus lylei* |
| EU603745 | Thailand | *Pteropus lylei* |
| EU603744 | Thailand | *Pteropus lylei* |
| EU603743 | Thailand | *Pteropus lylei* |
| EU603742 | Thailand | *Pteropus lylei* |
| EU603741 | Thailand | *Pteropus lylei* |
| EU603740 | Thailand | *Pteropus lylei* |
| EU603739 | Thailand | *Pteropus lylei* |
| EU603738 | Thailand | *Pteropus lylei* |
| EU603737 | Thailand | *Pteropus lylei* |
| EU603736 | Thailand | *Pteropus lylei* |
| EU603735 | Thailand | *Pteropus lylei* |
| EU603734 | Thailand | *Pteropus lylei* |
| EU603733 | Thailand | *Pteropus lylei* |
| EU603732 | Thailand | *Pteropus lylei* |
| EU603731 | Thailand | *Pteropus lylei* |
| EU603730 | Thailand | *Pteropus lylei* |
| EU603729 | Thailand | *Pteropus lylei* |
| EU603728 | Thailand | *Pteropus lylei* |
| EU603727 | Thailand | *Pteropus lylei* |
| EU603726 | Thailand | *Pteropus lylei* |
| EU603725 | Thailand | *Pteropus lylei* |
| EU603724 | Thailand | *Pteropus lylei* |
| EF070189 | Thailand | *Pteropus lylei* |
| EF070188 | Thailand | *Pteropus lylei* |
| EF070187 | Thailand | *Pteropus lylei* |
| EF070186 | Thailand | *Pteropus lylei* |
| EF070185 | Thailand | *Pteropus lylei* |
| EF070184 | Thailand | *Pteropus lylei* |
| EF070183 | Thailand | *Pteropus lylei* |
| EF070182 | Thailand | *Pteropus lylei* |
| JN808864 | Bangladesh | *Homo sapiens* |
| JN808863 | Bangladesh | *Homo sapiens* |
| JN808857 | Bangladesh | *Homo sapiens* |
| JN808860 | Bangladesh | *Homo sapiens* |
| JN808859 | Bangladesh | *Homo sapiens* |
| FJ513078 | India | *Homo sapiens* |
| FN869553 | Malaysia | *Pteropus vampyrus* |
| AY029767 | Malaysia | *Homo sapiens* |
| AY988601 | Bangladesh | *Homo sapiens* |
| AF376747 | Malaysia | *Pteropus hypomelanus* |
| AF212302 | Malaysia | *Homo sapiens* |
| AF238466 | Malaysia | *Homo sapiens* |
| AJ627196 | Malaysia | *Sus scrofa* |
| AJ564623 | Malaysia | *Homo sapiens* |
| AJ564622 | Malaysia | *Sus scrofa* |
| AJ564621 | Malaysia | *Sus scrofa* |
| MK575060 | Bangladesh | *Pteropus medius* |
| MK575061 | Bangladesh | *Pteropus medius* |
| MK575062 | Bangladesh | *Pteropus medius* |
| MK575063 | Bangladesh | *Pteropus medius* |
| MK575064 | Bangladesh | *Pteropus medius* |
| MK575065 | Bangladesh | *Pteropus medius* |
| MK575066 | Bangladesh | *Pteropus medius* |
| MK575067 | Bangladesh | *Pteropus medius* |
| MK575068 | Bangladesh | *Pteropus medius* |
| MK575069 | Bangladesh | *Pteropus medius* |
| MK575070 | Bangladesh | *Pteropus medius* |
| MT890728 | Bangladesh | *Homo sapiens* |
| MT890729 | Bangladesh | *Homo sapiens* |
| MT890711 | Bangladesh | *Homo sapiens* |
| MT890712 | Bangladesh | *Homo sapiens* |
| MT890714 | Bangladesh | *Homo sapiens* |
| MT890715 | Bangladesh | *Homo sapiens* |
| MT890719 | Bangladesh | *Homo sapiens* |
| MT890721 | Bangladesh | *Homo sapiens* |
| MT890723 | Bangladesh | *Homo sapiens* |
| MT890725 | Bangladesh | *Homo sapiens* |
| MT890726 | Bangladesh | *Homo sapiens* |
| MT890722 | Bangladesh | *Homo sapiens* |
| MT890724 | Bangladesh | *Homo sapiens* |
| MT890727 | Bangladesh | *Homo sapiens* |
| MT890730 | Bangladesh | *Homo sapiens* |
| MT890731 | Bangladesh | *Homo sapiens* |
| MT890720 | Bangladesh | *Homo sapiens* |
| MT890717 | Bangladesh | *Homo sapiens* |
| MT890716 | Bangladesh | *Homo sapiens* |
| MT890702 | Bangladesh | *Pteropus medius* |
| MT890703 | Bangladesh | *Pteropus medius* |
| MT890704 | Bangladesh | *Pteropus medius* |
| MT890705 | Bangladesh | *Pteropus medius* |
| MT890706 | Bangladesh | *Pteropus medius* |
| MT890707 | Bangladesh | *Pteropus medius* |
| MT890708 | Bangladesh | *Pteropus medius* |
| MT890709 | Bangladesh | *Pteropus medius* |
| MT890710 | Bangladesh | *Pteropus medius* |
| C011 | Cambodia | *Pteropus lylei* |
| C077 | Cambodia | *Pteropus lylei* |
| H001 | Cambodia | *Pteropus lylei* |
| B076 | Cambodia | *Pteropus lylei* |
| B079 | Cambodia | *Pteropus lylei* |
| FF099 | Cambodia | *Pteropus lylei* |
| FF107 | Cambodia | *Pteropus lylei* |
| FF031 | Cambodia | *Pteropus lylei* |
| FF044 | Cambodia | *Pteropus lylei* |
| FF047 | Cambodia | *Pteropus lylei* |
| FF048 | Cambodia | *Pteropus lylei* |
| KM034754 | Cambodia | *Pteropus lylei* |
| C282 | Cambodia | *Pteropus lylei* |
| Ba091 | Cambodia | *Pteropus lylei* |
| Ba097 | Cambodia | *Pteropus lylei* |
| Ba977 | Cambodia | *Pteropus lylei* |
| Ba996 | Cambodia | *Pteropus lylei* |
| Ba106 | Cambodia | *Pteropus lylei* |
| Ba108 | Cambodia | *Pteropus lylei* |
| Ba112 | Cambodia | *Pteropus lylei* |
| Ba132 | Cambodia | *Pteropus lylei* |
| Ba171 | Cambodia | *Pteropus lylei* |
| Ba173 | Cambodia | *Pteropus lylei* |
| Ba547 | Cambodia | *Pteropus lylei* |
| Ba555 | Cambodia | *Pteropus lylei* |
| Ba611 | Cambodia | *Pteropus lylei* |
| C271 | Cambodia | *Pteropus lylei* |
| KM034753 | Cambodia | *Pteropus lylei* |
| KM034755 | Cambodia | *Pteropus lylei* |
| MK995302 | Bangladesh | *Pteropus medius* |
| MK995301 | Bangladesh | *Pteropus medius* |
| MK995300 | Bangladesh | *Pteropus medius* |
| MK995299 | Bangladesh | *Pteropus medius* |
| MK995298 | Bangladesh | *Pteropus medius* |
| MK995295 | Bangladesh | *Pteropus medius* |
| MK995294 | Bangladesh | *Pteropus medius* |
| MK995293 | Bangladesh | *Pteropus medius* |
| MK995292 | Bangladesh | *Pteropus medius* |
| MK995291 | Bangladesh | *Pteropus medius* |
| MK995290 | Bangladesh | *Pteropus medius* |
| MK995289 | Bangladesh | *Pteropus medius* |
| MK995288 | Bangladesh | *Pteropus medius* |
| MK995287 | Bangladesh | *Pteropus medius* |
| MK995286 | Bangladesh | *Pteropus medius* |
| MK995285 | Bangladesh | *Pteropus medius* |
| MK995284 | Bangladesh | *Pteropus medius* |
| MK801755 | Cambodia | *Pteropus lylei* |
| MH423324 | India | *Homo sapiens* |
| MH423323 | India | *Homo sapiens* |
| MH523645 | India | *Pteropus medius* |
| MH523644 | India | *Pteropus medius* |
| MH523643 | India | *Pteropus medius* |
| MH523642 | India | *Homo sapiens* |
| MH523641 | India | *Homo sapiens* |
| MH523640 | India | *Homo sapiens* |
| MH891773 | India | *Homo sapiens* |
| MH396625 | India | *Homo sapiens* |
| MF133375 | Bangladesh | *Homo sapiens* |
| MW535746 | Thailand | *Pteropus lylei* |
| MW573860 | Thailand | *Pteropus lylei* |
| MW573861 | Thailand | *Pteropus lylei* |
| MW573862 | Thailand | *Pteropus lylei* |
| MW573863 | Thailand | *Pteropus lylei* |
| MW573864 | Thailand | *Pteropus lylei* |
| MW573865 | Thailand | *Pteropus lylei* |
| MW573866 | Thailand | *Pteropus lylei* |
| MW573867 | Thailand | *Pteropus lylei* |
| MW573868 | Thailand | *Pteropus lylei* |
| MW573869 | Thailand | *Pteropus lylei* |
| MW573870 | Thailand | *Pteropus lylei* |
| MW573871 | Thailand | *Pteropus lylei* |
| MW573872 | Thailand | *Pteropus lylei* |
| MW573873 | Thailand | *Pteropus lylei* |
| MW573874 | Thailand | *Pteropus lylei* |
| MW573875 | Thailand | *Pteropus lylei* |
| MW573876 | Thailand | *Pteropus lylei* |
| MW573877 | Thailand | *Pteropus lylei* |
| MW573878 | Thailand | *Pteropus lylei* |
| MW573879 | Thailand | *Pteropus lylei* |
| MW573880 | Thailand | *Pteropus lylei* |
| MK673558 | Malaysia | *Sus scrofa* |
| MK673559 | Malaysia | *Sus scrofa* |
| MK673560 | Malaysia | *Sus scrofa* |
| MK673561 | Malaysia | *Sus scrofa* |
| MK673562 | Malaysia | *Homo sapiens* |
| MK673563 | Malaysia | *Dog* |
| MK673565 | Bangladesh | *Homo sapiens* |
| MK673566 | Bangladesh | *Homo sapiens* |
| MK673567 | Bangladesh | *Homo sapiens* |
| MK673568 | Bangladesh | *Homo sapiens* |
| MK673570 | Bangladesh | *Homo sapiens* |
| MK673571 | Bangladesh | *Homo sapiens* |
| MK673572 | Bangladesh | *Homo sapiens* |
| MK673573 | Bangladesh | *Homo sapiens* |
| MK673574 | Bangladesh | *Homo sapiens* |
| MK673575 | Bangladesh | *Homo sapiens* |
| MK673576 | Bangladesh | *Homo sapiens* |
| MK673577 | Bangladesh | *Homo sapiens* |
| MK673578 | Bangladesh | *Homo sapiens* |
| MK673579 | Bangladesh | *Homo sapiens* |
| MK673580 | Bangladesh | *Homo sapiens* |
| MK673581 | Bangladesh | *Homo sapiens* |
| MK673582 | Bangladesh | *Homo sapiens* |
| MK673583 | Bangladesh | *Homo sapiens* |
| MK673584 | Bangladesh | *Homo sapiens* |
| MK673585 | Bangladesh | *Homo sapiens* |
| MK673586 | Bangladesh | *Homo sapiens* |
| MK673587 | Bangladesh | *Homo sapiens* |
| MK673588 | Bangladesh | *Homo sapiens* |
| MK673589 | Bangladesh | *Homo sapiens* |
| MK673590 | Bangladesh | *Homo sapiens* |
| MK673591 | Bangladesh | *Homo sapiens* |
| MK673592 | Bangladesh | *Homo sapiens* |

**Table S2:**
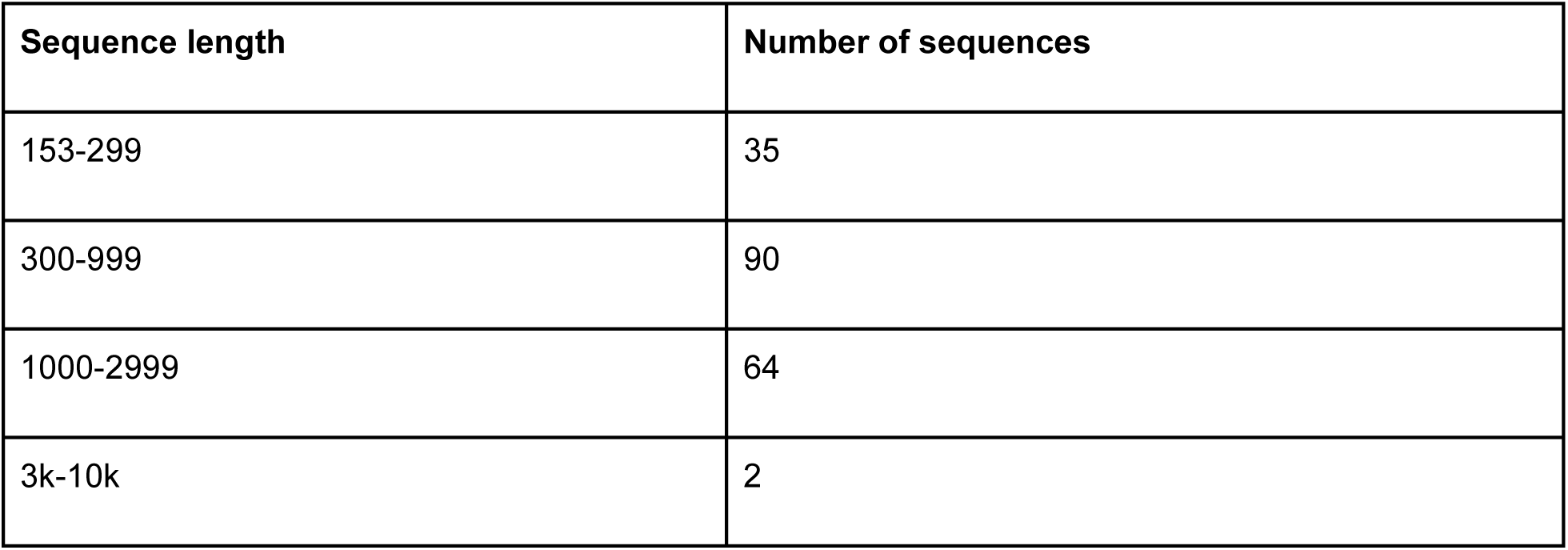
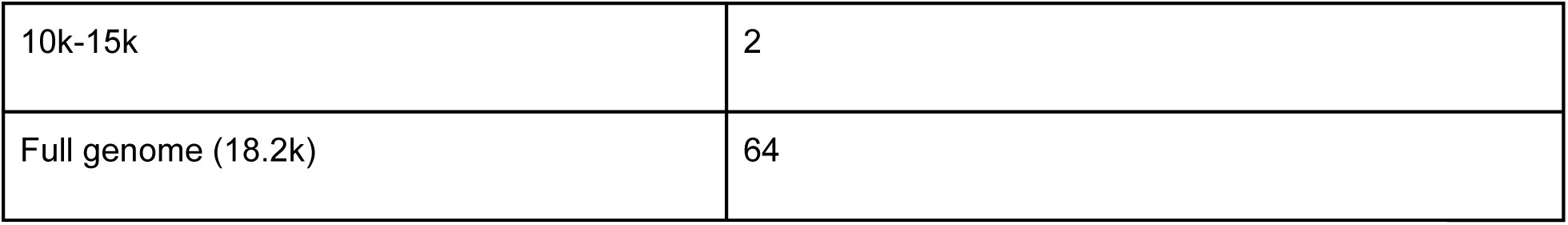
Number of sequences for different genome lengths.

| Sequence length | Number of sequences |
| --- | --- |
| 153-299 | 35 |
| 300-999 | 90 |
| 1000-2999 | 64 |
| 3k-10k | 2 |
| 10k-15k | 2 |
| Full genome (18.2k) | 64 |

**Table S3:** Input parameter values that resulted in at least 90% of tips on the reconstructed NiV phylogeny being assigned to a genetic cluster on PhyCLIP.

| **S** | **FDR** | **Gamma** | **Proportion of clustered sequences** |
| --- | --- | --- | --- |
| 3 | 0.25 | 3 | 0.947674 |
| 2 | 0.2 | 3 | 0.947674 |
| 2 | 0.25 | 3 | 0.947674 |
| 3 | 0.3 | 3 | 0.947674 |
| 1 | 0.3 | 3 | 0.947674 |
| 3 | 0.15 | 3 | 0.947674 |
| 1 | 0.25 | 3 | 0.947674 |
| 1 | 0.2 | 3 | 0.947674 |
| 3 | 0.1 | 3 | 0.947674 |
| 2 | 0.3 | 3 | 0.936047 |
| 2 | 0.1 | 3 | 0.930233 |
| 1 | 0.15 | 3 | 0.930233 |
| 3 | 0.2 | 3 | 0.924419 |
| 3 | 0.3 | 2 | 0.918605 |
| 3 | 0.25 | 2 | 0.912791 |
| 1 | 0.1 | 3 | 0.912791 |
| 2 | 0.15 | 3 | 0.906977 |
| 1 | 0.25 | 2 | 0.906977 |
| 1 | 0.2 | 2 | 0.906977 |
| 1 | 0.3 | 2 | 0.906977 |
| 2 | 0.15 | 2 | 0.901163 |
| 2 | 0.2 | 2 | 0.901163 |
| 3 | 0.1 | 2 | 0.901163 |

**Table S4:** Estimated asymptotic diversity for six subnational regions with more than 10 available sequences, as estimated by extrapolating Hill Numbers of order 0 with the iNEXT package on R, with several environmental characteristics (mean pairwise spatial distance, human population density, and percentage of tree cover) that were explored as potential ecological drivers of NiV diversity in South and Southeast Asia.

| Region | Estimated n. clusters | Mean Pairwise Spatial Distance (km) | Human Population Density (pop per km<sup>2</sup>) | Percentage Tree Cover |
| --- | --- | --- | --- | --- |
| BGD - Dhaka | 5 [5 - 6.2] | 31.0 | 2156 | 4.94 |
| BGD - Rajshahi | 3 [3 - 3] | 63.6 | 1121 | 0.26 |
| BGD - Rangpur | 2 [2 - 2.2] | 49.3 | 1088 | 0.46 |
| KHM - Kandal | 3 [3 - 3.4] | 0 | 378 | 15.8 |
| Central Thailand | 4 [4 - 5.3] | 64.3 | 221 | 17.0 |
| Eastern Thailand | 10 [7 - 13] | 14.6 | 142 | 32.9 |

**Table S5:** Hyperparameters and parameters used in our Approximate Bayesian Computation approach to estimate the spatial characteristics of NiV genetic clusters, with the values or prior distributions that we used to run our simulations.

| Type | Name | Description | Value used |
| --- | --- | --- | --- |
| Hyperparameter | $width_{outer}$ | Width of outer polygon (km) | 11000 |
| Hyperparameter | $width_{inner}$ | Width of inner polygon (km) | 5000 |
| Hyperparameter | $n_{obs}$ | Number of simulated samples per genetic cluster | 500 |
| Hyperparameter | $n_{sim}$ | Number of simulations | 10,000 |
| Parameter | $clust_{dens}$ | Cluster density per individual simulation | $\mathcal{U}([8.3 \times 10^{-9}, 2.9 \times 10^{-5}]))$ |
| Parameter | $sd_{clust}$ | Standard deviation of truncated normal distribution conditioning spatial spread of simulated clusters | $\mathcal{U}([1, 500]))$ |

**Table S6:** Estimations of the number of genetic clusters (with 95% Confidence Intervals in parentheses) for each country with *Pteropus* bats circulation, according to different estimates of *Pteropus* range (in km^2^): the IUCN’s estimations of *Pteropus* geographic range, and the Global Forest Watch’s estimations of tree cover per country above thresholds of 10%, 30%, and 50%, respectively. Countries where these data were not available are marked with a dash.

|  | IUCN *Pteropus* range |  | Tree cover (10% threshold) |  | Tree cover (30% threshold) |  | Tree cover (50% threshold) |  |
| --- | --- | --- | --- | --- | --- | --- | --- | --- |
| Country | Area | N. clusters | Area | N. clusters | Area | N. clusters | Area | N. clusters |
| American Samoa | 102 | 9 (3, 17) | - | - | - | - | - | - |
| Australia | 3,163,602 | 56 (24, 92) | 659,111 | 26 (11, 41) | 422,806 | 21 (9, 35) | 257,197 | 18 (8, 30) |
| Bangladesh | 127,346 | 15 (6, 25) | 26,607 | 11 (4, 20) | 19,390 | 11 (4, 20) | 15,229 | 10 (4, 19) |
| Bhutan | 25,222 | 11 (4, 20) | 26,792 | 11 (4, 20) | 25,772 | 11 (4, 20) | 23,665 | 11 (4, 20) |
| Brunei | 5,573 | 10 (3, 18) | 5,340 | 10 (3, 18) | 5,293 | 10 (3, 18) | 5,237 | 10 (3, 18) |
| Cambodia | 62,957 | 13 (5, 22) | 101,985 | 14 (6, 24) | 88,099 | 14 (6, 24) | 74,823 | 13 (5, 23) |
| Republic of China (Taiwan) | 10,957 | 10 (4, 19) | - | - | - | - | - | - |
| Comoros | 1,472 | 9 (3, 17) | 1,584 | 9 (3, 17) | 1,348 | 9 (3, 17) | 1,060 | 9 (3, 17) |
| Cook Is. | 49 | 9 (3, 17) | - | - | - | - | - | - |
| Fiji | 16,617 | 11 (4, 19) | 16,505 | 11 (4, 19) | 15,566 | 10 (4, 19) | 13,953 | 10 (4, 19) |
| France (Mayotte, Reunion) | 2,807 | 9 (3, 18) | 2,387 | 9 (3, 18) | 2,041 | 9 (3, 18) | 1620 | 9 (3, 17) |
| Guam | 451 | 9 (3, 17) | - | - | - | - | - | - |
| India | 2,987,747 | 55 (24, 89) | 490,910 | 23 (10, 37) | 388,304 | 21 (9, 34) | 304,653 | 19 (8, 31) |
| Indonesia | 1,656,164 | 40 (17, 63) | 1,650,987 | 39 (17, 63) | 1,606,412 | 39 (17, 62) | 1,550,634 | 38 (17, 61) |
| Japan (Ogasawara, Ryukyu, Iwo islands) | 2,082 | 9 (3, 17) | 7,957 | 10 (3, 19) | 7,654 | 10 (3, 19) | 7,341 | 10 (3, 18) |
| Madagascar | 185,931 | 16 (7, 27) | 331,909 | 20 (9, 32) | 171,411 | 16 (7, 27) | 122,414 | 15 (6, 25) |
| Malaysia | 323,929 | 20 (8, 32) | 297,861 | 19 (8, 31) | 294,306 | 19 (8, 31) | 289,772 | 19 (8, 31) |
| Maldives | 1 | 8 (3, 17) | 58 | 9 (3, 17) | 56 | 9 (3, 17) | 45 | 9 (3, 17) |
| Mauritius | 1,805 | 9 (3, 17) | 1,487 | 9 (3, 17) | 713 | 9 (3, 17) | 509 | 9 (3, 17) |
| Micronesia | 351 | 9 (3, 17) | 65 | 9 (3, 17) | 62 | 9 (3, 17) | 60 | 9 (3, 17) |
| Myanmar | 324,734 | 20 (8, 32) | 450,853 | 22 (10, 35) | 428,634 | 22 (9, 35) | 397,937 | 21 (9, 34) |
| N. Mariana Is. | 314 | 9 (3, 17) | - | - | - | - | - | - |
| Nepal | 99,446 | 14 (6, 24) | 56,000 | 13 (5, 22) | 51,610 | 12 (5, 22) | 43,082 | 12 (5, 21) |
| New Caledonia | 17,834 | 11 (4, 19) | 15,978 | 10 (4, 19) | 14,403 | 10 (4, 19) | 10,513 | 10 (4, 19) |
| Niue | 192 | 9 (3, 17) | - | - | - | - | - | - |
| Pakistan | 400,735 | 21 (9, 34) | 16,299 | 11 (4, 19) | 9,787 | 10 (4, 19) | 6,669 | 10 (3, 18) |
| Papua New Guinea | 370,664 | 20 (9, 33) | 436,905 | 22 (10, 35) | 429,218 | 22 (9, 35) | 419,768 | 21 (9, 35) |
| Philippines | 275,600 | 19 (8, 30) | 197,985 | 17 (7, 28) | 185,994 | 16 (7, 28) | 174,152 | 16 (7, 27) |
| Samoa | 2,567 | 9 (3, 18) | - | - | - | - | - | - |
| Seychelles | 104 | 9 (3, 17) | 126 | 9 (3, 17) | 62 | 9 (3, 17) | 29 | 8 (3, 17) |
| Singapore | 457 | 9 (3, 17) | 218 | 9 (3, 17) | 188 | 9 (3, 17) | 168 | 9 (3, 17) |
| Solomon Islands | 23,733 | 11 (4, 20) | 27,502 | 11 (4, 20) | 27,416 | 11 (4, 20) | 27,318 | 11 (4, 20) |
| Sri Lanka | 65,258 | 13 (5, 23) | 42,412 | 12 (5, 21) | 39,399 | 12 (4, 21) | 35,460 | 12 (4, 21) |
| Tanzania (Pemba, Mafia islands) | 1,179 | 9 (3, 17) | 1,344 | 9 (3, 17) | 909 | 9 (3, 17) | 683 | 9 (3, 17) |
| Thailand | 138,538 | 15 (6, 26) | 219,195 | 17 (7, 29) | 199,624 | 17 (7, 28) | 179,669 | 16 (7, 27) |
| Timor-Leste | 14,405 | 10 (4, 19) | 8,441 | 10 (3, 19) | 7,262 | 10 (3, 18) | 6,169 | 10 (3, 18) |
| Tonga | 343 | 9 (3, 17) | - | - | - | - | - | - |
| Vanuatu | 10,791 | 10 (4, 19) | 11,861 | 10 (4, 19) | 11,812 | 10 (4, 19) | 11,615 | 10 (4, 19) |
| Vietnam | 36,012 | 12 (4, 21) | 181,462 | 16 (7, 27) | 165,513 | 16 (7, 27) | 147,599 | 15 (6, 26) |
| Wallis and Futuna Is. | 86 | 9 (3, 17) | - | - | - | - | - | - |
| **Total** | **10,358,157** | **118 (49, 211)** | **5,288,126** | **76 (32, 129)** | **4,621,064** | **70 (30, 118)** | **4,129,042** | **66 (28, 109)** |

## References

1. World Health Organization, NIPAH RESEARCH AND DEVELOPMENT (R&D) ROADMAP (2019) (available at https://cdn.who.int/media/docs/default-source/blue-print/nipah_rdblueprint_roadmap_advanceddraftoct2019.pdf?sfvrsn=4f0dc9ad_3&download=true).

2. B. Nikolay, H. Salje, M. J. Hossain, A. K. M. D. Khan, H. M. S. Sazzad, M. Rahman, P. Daszak, U. Ströher, J. R. C. Pulliam, A. M. Kilpatrick, S. T. Nichol, J. D. Klena, S. Sultana, S. Afroj, S. P. Luby, S. Cauchemez, E. S. Gurley, Transmission of Nipah Virus - 14 Years of Investigations in Bangladesh. N. Engl. J. Med. 380, 1804–1814 (2019).

3. R. K. Plowright, D. J. Becker, D. E. Crowley, A. D. Washburne, T. Huang, P. O. Nameer, E. S. Gurley, B. A. Han, Prioritizing surveillance of Nipah virus in India. PLoS Negl. Trop. Dis. 13, e0007393 (2019).

4. V. Soman Pillai, G. Krishna, M. Valiya Veettil, Nipah Virus: Past Outbreaks and Future Containment. Viruses 12 (2020), doi:10.3390/v12040465.

5. M. K. Lo, L. Lowe, K. B. Hummel, H. M. S. Sazzad, E. S. Gurley, M. J. Hossain, S. P. Luby, D. M. Miller, J. A. Comer, P. E. Rollin, W. J. Bellini, P. A. Rota, Characterization of Nipah virus from outbreaks in Bangladesh, 2008-2010. Emerg. Infect. Dis. 18, 248–255 (2012).

6. K. Halpin, A. D. Hyatt, R. Fogarty, D. Middleton, J. Bingham, J. H. Epstein, S. A. Rahman, T. Hughes, C. Smith, H. E. Field, P. Daszak, Henipavirus Ecology Research Group, Pteropid bats are confirmed as the reservoir hosts of henipaviruses: a comprehensive experimental study of virus transmission. Am. J. Trop. Med. Hyg. 85, 946–951 (2011).

7. P. K. G. Ching, V. C. de los Reyes, M. N. Sucaldito, E. Tayag, A. B. Columna-Vingno, F. F. Malbas Jr, G. C. Bolo Jr, J. J. Sejvar, D. Eagles, G. Playford, E. Dueger, Y. Kaku, S. Morikawa, M. Kuroda, G. A. Marsh, S. McCullough, A. R. Foxwell, Outbreak of henipavirus infection, Philippines, 2014. Emerg. Infect. Dis. 21, 328–331 (2015).

8. C. D. McKee, A. Islam, S. P. Luby, H. Salje, P. J. Hudson, R. K. Plowright, E. S. Gurley, The Ecology of Nipah Virus in Bangladesh: A Nexus of Land-Use Change and Opportunistic Feeding Behavior in Bats. Viruses 13 (2021), doi:10.3390/v13020169.

9. J. H. Epstein, S. J. Anthony, A. Islam, A. M. Kilpatrick, S. Ali Khan, M. D. Balkey, N. Ross, I. Smith, C. Zambrana-Torrelio, Y. Tao, A. Islam, P. L. Quan, K. J. Olival, M. S. U. Khan, E. S. Gurley, M. J. Hossein, H. E. Field, M. D. Fielder, T. Briese, M. Rahman, C. C. Broder, G. Crameri, L.-F. Wang, S. P. Luby, W. I. Lipkin, P. Daszak, Nipah virus dynamics in bats and implications for spillover to humans. Proc. Natl. Acad. Sci. U. S. A. 117, 29190–29201 (2020).

10. S. T. Hegde, H. Salje, H. M. S. Sazzad, M. J. Hossain, M. Rahman, P. Daszak, J. D. Klena, S. T. Nichol, S. P. Luby, E. S. Gurley, Using healthcare-seeking behaviour to estimate the number of Nipah outbreaks missed by hospital-based surveillance in Bangladesh. Int. J. Epidemiol. 48, 1219–1227 (2019).

11. J.-M. Reynes, D. Counor, S. Ong, C. Faure, V. Seng, S. Molia, J. Walston, M. C. Georges-Courbot, V. Deubel, J.-L. Sarthou, Nipah virus in Lyle’s flying foxes, Cambodia. Emerg. Infect. Dis. 11, 1042–1047 (2005).

12. M. Z. Rahman, M. M. Islam, M. E. Hossain, M. M. Rahman, A. Islam, A. Siddika, M. S. S. Hossain, S. Sultana, A. Islam, M. Rahman, M. Rahman, J. D. Klena, M. S. Flora, P. Daszak, J. H. Epstein, S. P. Luby, E. S. Gurley, Genetic diversity of Nipah virus in Bangladesh. Int. J. Infect. Dis. 102, 144–151 (2021).

13. J. Cappelle, T. Hoem, V. Hul, N. Furey, K. Nguon, S. Prigent, L. Dupon, S. Ken, C. Neung, V. Hok, L. Pring, T. Lim, S. Bumrungsri, R. Duboz, P. Buchy, S. Ly, V. Duong, A. Tarantola, A. Binot, P. Dussart, Nipah virus circulation at human-bat interfaces, Cambodia. Bull. World Health Organ. 98, 539–547 (2020).

14. S. Duchene, P. Lemey, T. Stadler, S. Y. W. Ho, D. A. Duchene, V. Dhanasekaran, G. Baele, Bayesian Evaluation of Temporal Signal in Measurably Evolving Populations. Mol. Biol. Evol. 37, 3363–3379 (2020).

15. A. Lo Presti, E. Cella, M. Giovanetti, A. Lai, S. Angeletti, G. Zehender, M. Ciccozzi, Origin and evolution of Nipah virus. J. Med. Virol. 88, 380–388 (2016).

16. A. X. Han, E. Parker, F. Scholer, S. Maurer-Stroh, C. A. Russell, Phylogenetic Clustering by Linear Integer Programming (PhyCLIP). Mol. Biol. Evol. 36, 1580–1595 (2019).

17. L. Hubert, P. Arabie, Comparing partitions. J. Classification 2, 193–218 (1985).

18. IUCN (International Union for Conservation of Nature), The IUCN Red List of Threatened Species. Version 2022-2IUCN (2022) (available at https://www.iucnredlist.org).

19. M. C. Hansen, P. V. Potapov, R. Moore, M. Hancher, S. A. Turubanova, A. Tyukavina, D. Thau, S. V. Stehman, S. J. Goetz, T. R. Loveland, A. Kommareddy, A. Egorov, L. Chini, C. O. Justice, J. R. G. Townshend, High-resolution global maps of 21st-century forest cover change. Science 342, 850–853 (2013).

20. Global Administrative Areas Database, version 3.6 (available at http://gadm.org/).

21. S. L. M. Whitmer, M. K. Lo, H. M. S. Sazzad, S. Zufan, E. S. Gurley, S. Sultana, B. Amman, J. T. Ladner, M. Z. Rahman, S. Doan, S. M. Satter, M. S. Flora, J. M. Montgomery, S. T. Nichol, C. F. Spiropoulou, J. D. Klena, Inference of Nipah virus evolution, 1999-2015. Virus Evol 7, veaa062 (2021).

22. M. B. Hahn, J. H. Epstein, E. S. Gurley, M. S. Islam, S. P. Luby, P. Daszak, J. A. Patz, Roosting behaviour and habitat selection of Pteropus giganteus reveals potential links to Nipah virus epidemiology. J. Appl. Ecol. 51, 376–387 (2014).

23. A. Chaiyes, P. Duengkae, S. Wacharapluesadee, N. Pongpattananurak, K. J. Olival, T. Hemachudha, Assessing the distribution, roosting site characteristics, and population of Pteropus lylei in Thailand. Raffles Bull. Zool. 65, 670–680 (2017).

24. S. Ravon, N. M. Furey, H. U. L. Vibol, J. Cappelle, A rapid assessment of fl ying fox (Pteropus spp.) colonies in Cambodia (available at http://www.seabcru.org/wp-content/uploads/2014/09/Ravon-et-al.-2014.-Cambodian-Pteropus.pdf).

25. E. E. Glennon, D. J. Becker, A. J. Peel, R. Garnier, R. D. Suu-Ire, L. Gibson, D. T. S. Hayman, J. L. N. Wood, A. A. Cunningham, R. K. Plowright, O. Restif, What is stirring in the reservoir? Modelling mechanisms of henipavirus circulation in fruit bat hosts. Philos. Trans. R. Soc. Lond. B Biol. Sci. 374, 20190021 (2019).

26. S. B. Kasloff, A. Leung, B. S. Pickering, G. Smith, E. Moffat, B. Collignon, C. Embury-Hyatt, D. Kobasa, H. M. Weingartl, Pathogenicity of Nipah henipavirus Bangladesh in a swine host. Sci. Rep. 9, 5230 (2019).

27. S. Kenmoe, M. Demanou, J. J. Bigna, C. Nde Kengne, A. Fatawou Modiyinji, F. B. N. Simo, S. Eyangoh, S. A. Sadeuh-Mba, R. Njouom, Case fatality rate and risk factors for Nipah virus encephalitis: A systematic review and meta-analysis. J. Clin. Virol. 117, 19–26 (2019).

28. E. S. Gurley, S. T. Hegde, K. Hossain, H. M. S. Sazzad, M. J. Hossain, M. Rahman, M. A. Y. Sharker, H. Salje, M. S. Islam, J. H. Epstein, S. U. Khan, A. M. Kilpatrick, P. Daszak, S. P. Luby, Convergence of Humans, Bats, Trees, and Culture in Nipah Virus Transmission, Bangladesh. Emerg. Infect. Dis. 23, 1446–1453 (2017).

29. E. de Wit, V. J. Munster, Animal models of disease shed light on Nipah virus pathogenesis and transmission. J. Pathol. 235, 196–205 (2015).

30. M. Gaudino, N. Aurine, C. Dumont, J. Fouret, M. Ferren, C. Mathieu, O. Reynard, V. E. Volchkov, C. Legras-Lachuer, M.-C. Georges-Courbot, B. Horvat, High Pathogenicity of Nipah Virus from Pteropus lylei Fruit Bats, Cambodia. Emerg. Infect. Dis. 26, 104–113 (2020).

31. S. Hegde, K. H. Lee, A. Styczynski, F. Jones, I. Gomes, P. Das, E. S. Gurley, Potential for person-to-person transmission of henipaviruses: A systematic review of the literaturebioRxiv (2023), doi:10.1101/2023.02.26.23286473.

32. M. C. Cortes, S. Cauchemez, N. Lefrancq, S. P. Luby, M. Jahangir Hossain, H. M. S. Sazzad, M. Rahman, P. Daszak, H. Salje, E. S. Gurley, Characterization of the Spatial and Temporal Distribution of Nipah Virus Spillover Events in Bangladesh, 2007–2013. J. Infect. Dis. 217, 1390–1394 (2018).

33. E. J. Annand, B. A. Horsburgh, K. Xu, P. A. Reid, B. Poole, M. C. de Kantzow, N. Brown, A. Tweedie, M. Michie, J. D. Grewar, A. E. Jackson, N. B. Singanallur, K. M. Plain, K. Kim, M. Tachedjian, B. van der Heide, S. Crameri, D. T. Williams, C. Secombe, E. D. Laing, S. Sterling, L. Yan, L. Jackson, C. Jones, R. K. Plowright, A. J. Peel, A. C. Breed, I. Diallo, N. K. Dhand, P. N. Britton, C. C. Broder, I. Smith, J.-S. Eden, Novel Hendra Virus Variant Detected by Sentinel Surveillance of Horses in Australia. Emerg. Infect. Dis. 28, 693–704 (2022).

34. K. Clark, I. Karsch-Mizrachi, D. J. Lipman, J. Ostell, E. W. Sayers, GenBank. Nucleic Acids Res. 44, D67–72 (2016).

35. S. Kumar, G. Stecher, M. Li, C. Knyaz, K. Tamura, MEGA X: Molecular Evolutionary Genetics Analysis across Computing Platforms. Mol. Biol. Evol. 35, 1547–1549 (2018).

36. K. B. Chua, W. J. Bellini, P. A. Rota, B. H. Harcourt, A. Tamin, S. K. Lam, T. G. Ksiazek, P. E. Rollin, S. R. Zaki, W. Shieh, C. S. Goldsmith, D. J. Gubler, J. T. Roehrig, B. Eaton, A. R. Gould, J. Olson, H. Field, P. Daniels, A. E. Ling, C. J. Peters, L. J. Anderson, B. W. Mahy, Nipah virus: a recently emergent deadly paramyxovirus. Science 288, 1432–1435 (2000).

37. B. H. Harcourt, A. Tamin, T. G. Ksiazek, P. E. Rollin, L. J. Anderson, W. J. Bellini, P. A. Rota, Molecular characterization of Nipah virus, a newly emergent paramyxovirus. Virology 271, 334– 349 (2000).

38. Y. P. Chan, K. B. Chua, C. L. Koh, M. E. Lim, S. K. Lam, Complete nucleotide sequences of Nipah virus isolates from Malaysia. J. Gen. Virol. 82, 2151–2155 (2001).

39. K. B. Chua, C. L. Koh, P. S. Hooi, K. F. Wee, J. H. Khong, B. H. Chua, Y. P. Chan, M. E. Lim, S. K. Lam, Isolation of Nipah virus from Malaysian Island flying-foxes. Microbes Infect. 4, 145– 151 (2002).

40. S. AbuBakar, L.-Y. Chang, A. R. M. Ali, S. H. Sharifah, K. Yusoff, Z. Zamrod, Isolation and molecular identification of Nipah virus from pigs. Emerg. Infect. Dis. 10, 2228–2230 (2004).

41. B. H. Harcourt, L. Lowe, A. Tamin, X. Liu, B. Bankamp, N. Bowden, P. E. Rollin, J. A. Comer, T. G. Ksiazek, M. J. Hossain, E. S. Gurley, R. F. Breiman, W. J. Bellini, P. A. Rota, Genetic characterization of Nipah virus, Bangladesh, 2004. Emerg. Infect. Dis. 11, 1594–1597 (2005).

42. S. Wacharapluesadee, T. Hemachudha, Duplex nested RT-PCR for detection of Nipah virus RNA from urine specimens of bats. J. Virol. Methods 141, 97–101 (2007).

43. M. K. Lo, B. H. Harcourt, B. A. Mungall, A. Tamin, M. E. Peeples, W. J. Bellini, P. A. Rota, Determination of the henipavirus phosphoprotein gene mRNA editing frequencies and detection of the C, V and W proteins of Nipah virus in virus-infected cells. J. Gen. Virol. 90, 398–404 (2009).

44. S. Wacharapluesadee, K. Boongird, S. Wanghongsa, N. Ratanasetyuth, P. Supavonwong, D. Saengsen, G. N. Gongal, T. Hemachudha, A longitudinal study of the prevalence of Nipah virus in Pteropus lylei bats in Thailand: evidence for seasonal preference in disease transmission. Vector Borne Zoonotic Dis. 10, 183–190 (2010).

45. S. A. Rahman, S. S. Hassan, K. J. Olival, M. Mohamed, L.-Y. Chang, L. Hassan, N. M. Saad, S. A. Shohaimi, Z. C. Mamat, M. S. Naim, J. H. Epstein, A. S. Suri, H. E. Field, P. Daszak, Henipavirus Ecology Research Group, Characterization of Nipah virus from naturally infected Pteropus vampyrus bats, Malaysia. Emerg. Infect. Dis. 16, 1990–1993 (2010).

46. V. A. Arankalle, B. T. Bandyopadhyay, A. Y. Ramdasi, R. Jadi, D. R. Patil, M. Rahman, M. Majumdar, P. S. Banerjee, A. K. Hati, R. P. Goswami, D. K. Neogi, A. C. Mishra, Genomic characterization of Nipah virus, West Bengal, India. Emerg. Infect. Dis. 17, 907–909 (2011).

47. P. D. Yadav, C. G. Raut, A. M. Shete, A. C. Mishra, J. S. Towner, S. T. Nichol, D. T. Mourya, Detection of Nipah virus RNA in fruit bat (Pteropus giganteus) from India. Am. J. Trop. Med. Hyg. 87, 576–578 (2012).

48. I. Sendow, A. Ratnawati, T. Taylor, R. M. A. Adjid, M. Saepulloh, J. Barr, F. Wong, P. Daniels, H. Field, Nipah virus in the fruit bat Pteropus vampyrus in Sumatera, Indonesia. PLoS One 8, e69544 (2013).

49. S. Wacharapluesadee, P. Samseeneam, M. Phermpool, T. Kaewpom, A. Rodpan, P. Maneeorn, P. Srongmongkol, B. Kanchanasaka, T. Hemachudha, Molecular characterization of Nipah virus from Pteropus hypomelanus in Southern Thailand. Virol. J. 13, 53 (2016).

50. M. Z. Hassan, H. M. S. Sazzad, S. P. Luby, K. Sturm-Ramirez, M. U. Bhuiyan, M. Z. Rahman, M. M. Islam, U. Ströher, S. Sultana, M. A. H. Kafi, P. Daszak, M. Rahman, E. S. Gurley, Nipah Virus Contamination of Hospital Surfaces during Outbreaks, Bangladesh, 2013-2014. Emerg. Infect. Dis. 24, 15–21 (2018).

51. G. Arunkumar, R. Chandni, D. T. Mourya, S. K. Singh, R. Sadanandan, P. Sudan, B. Bhargava, Nipah Investigators People and Health Study Group, Outbreak Investigation of Nipah Virus Disease in Kerala, India, 2018. J. Infect. Dis. 219, 1867–1878 (2019).

52. D. E. Anderson, A. Islam, G. Crameri, S. Todd, A. Islam, S. U. Khan, A. Foord, M. Z. Rahman, I. H. Mendenhall, S. P. Luby, E. S. Gurley, P. Daszak, J. H. Epstein, L.-F. Wang, Isolation and Full-Genome Characterization of Nipah Viruses from Bats, Bangladesh. Emerg. Infect. Dis. 25, 166–170 (2019).

53. K. J. Olival, A. Latinne, A. Islam, J. H. Epstein, R. Hersch, R. C. Engstrand, E. S. Gurley, G. Amato, S. P. Luby, P. Daszak, Population genetics of fruit bat reservoir informs the dynamics, distribution and diversity of Nipah virus. Mol. Ecol. 29, 970–985 (2020).

54. S. Wacharapluesadee, S. Ghai, P. Duengkae, P. Manee-Orn, W. Thanapongtharm, A. W. Saraya, S. Yingsakmongkon, Y. Joyjinda, S. Suradhat, W. Ampoot, B. Nuansrichay, T. Kaewpom, R. Tantilertcharoen, A. Rodpan, K. Wongsathapornchai, T. Ponpinit, R. Buathong, S. Bunprakob, S. Damrongwatanapokin, C. Ruchiseesarod, S. Petcharat, W. Kalpravidh, K. J. Olival, M. M. Stokes, T. Hemachudha, Two decades of one health surveillance of Nipah virus in Thailand. One Health Outlook 3, 12 (2021).

55. S. Wacharapluesadee, T. Ngamprasertwong, T. Kaewpom, P. Kattong, A. Rodpan, S. Wanghongsa, T. Hemachudha, Genetic characterization of Nipah virus from Thai fruit bats (Pteropus lylei). Asian Biomed. 7, 813–819 (2013).

56. S. Kalyaanamoorthy, B. Q. Minh, T. K. F. Wong, A. von Haeseler, L. S. Jermiin, ModelFinder: fast model selection for accurate phylogenetic estimates. Nat. Methods 14, 587–589 (2017).

57. G. Baele, P. Lemey, T. Bedford, A. Rambaut, M. A. Suchard, A. V. Alekseyenko, Improving the accuracy of demographic and molecular clock model comparison while accommodating phylogenetic uncertainty. Mol. Biol. Evol. 29, 2157–2167 (2012).

58. G. Baele, W. L. S. Li, A. J. Drummond, M. A. Suchard, P. Lemey, Accurate model selection of relaxed molecular clocks in bayesian phylogenetics. Mol. Biol. Evol. 30, 239–243 (2013).

59. M. A. Suchard, P. Lemey, G. Baele, D. L. Ayres, A. J. Drummond, A. Rambaut, Bayesian phylogenetic and phylodynamic data integration using BEAST 1.10. Virus Evol 4, vey016 (2018).

60. V. Hill, G. Baele, Bayesian estimation of past population dynamics in BEAST 1.10 using the Skygrid coalescent model. Mol. Biol. Evol. (2019), doi:10.1093/molbev/msz172.

61. Rambaut, Drummond, LogCombiner v1. 8.2. LogCombinerv1 (2015).

62. A. Rambaut, A. J. Drummond, D. Xie, G. Baele, M. A. Suchard, Posterior Summarization in Bayesian Phylogenetics Using Tracer 1.7. Syst. Biol. 67, 901–904 (2018).

63. A. Rambaut, D. Aj, TreeAnnotator v. 2.3. 0. Part of the BEAST package(2014).

64. Rambaut, FigTree v1. 3.1. http://tree.bio.ed.ac.uk/software/figtree/ (2009) (available at https://cir.nii.ac.jp/crid/1572824500617143552).

65. G. Yu, D. K. Smith, H. Zhu, Y. Guan, T. T.-Y. Lam, Ggtree: An r package for visualization and annotation of phylogenetic trees with their covariates and other associated data. Methods Ecol. Evol. 8, 28–36 (2017).

66. G. Yu, T. T.-Y. Lam, H. Zhu, Y. Guan, Two methods for mapping and visualizing associated data on phylogeny using ggtreeMolecular Biology and Evolution 35, 3041–3043 (2018).

67. G. Yu, Using ggtree to Visualize Data on Tree-Like Structures Current Protocols in Bioinformatics 69, e96 (2020).

68. 68. G. Yu, Data Integration, Manipulation and Visualization of Phylogenetic Treess (2022) (available at https://www.amazon.com/Integration-Manipulation-Visualization-Phylogenetic-Computational-ebook/dp/B0B5NLZR1Z/).

69. 69. R Core Team, R: A Language and Environment for Statistical Computing (2022) (available at https://www.R-project.org/).

70. A. Chakraborty, H. M. S. Sazzad, M. J. Hossain, M. S. Islam, S. Parveen, M. Husain, S. S. Banu, G. Podder, S. Afroj, P. E. Rollin, P. Daszak, S. P. Luby, M. Rahman, E. S. Gurley, Evolving epidemiology of Nipah virus infection in Bangladesh: evidence from outbreaks during 2010-2011. Epidemiol. Infect. 144, 371–380 (2016).

71. M. Padgham, geodist: Fast, Dependency-Free Geodesic Distance Calculations (Github, 2021; https://github.com/hypertidy/geodist).

72. E. Paradis, K. Schliep, ape 5.0: an environment for modern phylogenetics and evolutionary analyses in R. Bioinformatics 35, 526–528 (2019).

73. M. J. Vavrek, fossil: palaeoecological and palaeogeographical analysis toolsPalaeontologia Electronica 14, 1T (2011).

74. T. C. Hsieh, K. H. Ma, A. Chao, iNEXT: an R package for rarefaction and extrapolation of species diversity (Hill numbers). Methods Ecol. Evol. 7, 1451–1456 (2016).

75. A. Chao, N. J. Gotelli, T. C. Hsieh, E. L. Sander, K. H. Ma, R. K. Colwell, A. M. Ellison, Rarefaction and extrapolation with Hill numbers: a framework for sampling and estimation in species diversity studies. Ecol. Monogr. 84, 45–67 (2014).

76. Bangladesh Bureau of Statistics, Ministry of Planning, Population and Housing Census 2022 (2022).

77. National Institute of Statistics, Ministry of Planning, General Population Census of the Kingdom of Cambodia 2019 (2020; http://nis.gov.kh/nis/Census2019/Final%20General%20Population%20Census%202019-English.pdf).

78. United Nations Development Programme, Advancing Human Development through the ASEAN Community, Thailand Human Development Report 2014 (2014; https://planipolis.iiep.unesco.org/sites/default/files/ressources/thailand_nhdr_2014_0.pdf).

79. K. Csilléry, O. François, M. G. B. Blum, abc: an R package for approximate Bayesian computation (ABC). Methods Ecol. Evol. 3, 475–479 (2012).

80. G. Van Rossum, F. L. Drake, Python 3 Reference Manual: (Python Documentation Manual Part 2) (CreateSpace Independent Publishing Platform, 2009; https://play.google.com/store/books/details?id=KIybQQAACAAJ).

81. F. Pedregosa, G. Varoquaux, A. Gramfort, V. Michel, B. Thirion, O. Grisel, M. Blondel, P. Prettenhofer, R. Weiss, V. Dubourg, J. Vanderplas, A. Passos, D. Cournapeau, M. Brucher, M. Perrot, E. Duchesnay, Scikit-learn: Machine Learning in Python. J. Mach. Learn. Res.

